# Modeling nonlinear and interaction effects of spatiotemporal and other non-genetic factors improves phenotypic prediction for complex traits

**DOI:** 10.1101/2025.11.26.25341090

**Authors:** Ross DeVito, Melissa Gymrek

## Abstract

Adjusting for non-genetic factors can improve genetic association testing and polygenic prediction, yet most studies rely on linear adjustments for a small set of covariates. Location and time covariates are typically not included in genetic studies, but they can serve as useful proxies for environmental exposures such as climate, pollution, and behavioural differences. These features especially highlight the weakness of linear covariate adjustments, which cannot capture their cyclical or nonlinear effects nor interactions between covariates. To evaluate the benefits of modeling nonlinear covariate effects, including spatiotemporal features, we adopted a null model approach in which an auxiliary nonlinear model predicts the phenotype from non-genetic covariates alone. This prediction is then included as an additional covariate in downstream analysis, allowing association studies, polygenic scores, and fine-mapping to account for nonlinear covariate effects and interactions without modifying existing tools. Using 16 continuous phenotypes from the UK Biobank, we first evaluated gradient boosted decision tree (GBDT) and neural network null models with combinations of additional time-of-day, time-of-year, and birth or home location covariates and determined how well these models could predict phenotypes from covariates alone compared to linear models. A GBDT null improved average R^2^ by 4.3% over a linear null across all phenotype-covariate set pairs, and models including spatiotemporal covariates achieved the best performance for all phenotypes. Incorporating nonlinear null predictions with additional spatiotemporal features improved polygenic scores for all phenotypes, with median improvements of 7.3% for BASIL and 15.5% for PRS-CS over the standard approach. We additionally tested three case-control phenotypes and observed improvement beyond the 95% CI of the standard approach for depression. Finally, model interpretation identified both known and novel covariate interactions and nonlinear effects. This highlighted several important cases which linear adjustments would be unable to capture, including examples where phenotype means exhibit opposite age-related trends between sexes, cyclical temporal patterns for vitamin D reflecting seasonal variation, and complex spatial dependencies between birth and home locations. Together, these results demonstrate that a small addition to existing genetic analysis workflows can improve predictive performance and provide new insights on environmental effects for complex traits.

## Introduction

Statistical methods for understanding the genetic basis of complex traits, such as genome wide association studies (GWAS), polygenic scores (PGS), and fine-mapping, rely on identifying associations between genetic variants and phenotypes. Adjusting for other sources of phenotypic variation with covariates in these models is critical to improve statistical power and reduce confounding (1–3). Common covariates include age and sex, which explain a substantial portion of variability for many phenotypes, as well as genotype principal components (PCs), which help control for confounding due to population structure or shared ancestry (1,3–6). These covariates may also serve as proxies for unmeasured sources of variation, such as cultural or lifestyle differences (6).

Standard approaches for modeling covariates face multiple limitations. First, covariates are typically added as additional terms to linear models, which scale efficiently to handle large-scale data. However, this approach assumes that covariates have linear effects and do not interact with each other or with genetic variants included in the model. Second, although these models are simple and straightforward to interpret, violation of the linear model assumption can make model interpretation unreliable (7). Third, most analyses rely on a limited set of covariates (e.g. age, sex, PCs), ignoring other features such as spatiotemporal information with potentially large impacts on phenotypes.

DeepNull (5) addressed the first of these limitations by modeling nonlinear and interaction effects among covariates. In their framework, a deep learning model is used to predict a phenotype from covariates (age, sex, genotyping array) alone. The prediction from this auxiliary model, which they refer to as a null model, is then used as a single covariate in downstream linear models. This two-stage process effectively removes complex effects of covariates not captured by standard models while remaining compatible with linear association testing implemented by existing tools. Their study showed this approach improved statistical power in GWAS simulations, increased the number of significant signals in real phenotypes, and improved PGS performance. While the DeepNull approach can in theory also address the second challenge of interpretation by overcoming model misspecification imposed by linear models, the model types used (deep neural networks) can be difficult to reliably interpret (8–11). Whereas DeepNull focused solely on modeling covariate effects, alternative strategies have been developed that perform similar modeling of nonlinear effects and interactions across both genotype and covariate features (12–14). However, those approaches necessitate aggressive feature selection to reduce the number of genetic variants considered in order to make training feasible, making it possible that truly causal variants will be excluded from the final model. As a result, we focus on covariate modeling here, though the null model approach could be integrated into their feature selection process.

The third challenge is which additional covariates to include. The set of covariates included is critical regardless of the modeling approach used. DeepNull, like most standard GWAS, relied on a small number of commonly used covariates (e.g. age and sex). While some environmental covariates are well-known to influence certain traits (e.g. sun exposure for vitamin D (15) or physical activity for heart disease (16)), these features are often not readily available and it can be cumbersome to manually identify relevant covariates for each trait. Instead, we sought to identify a set of general-purpose covariates capable of acting as proxies for a wide array of environmental factors. This led us to focus on spatiotemporal features: birth and home locations and time of both year and day of assessment. These covariates are typically available in large cohorts and are broadly relevant across many traits, as they reflect spatial and temporal variation in environmental factors such as climate, pollution, and behavior. Importantly, their effects are unlikely to be well captured by linear models. For example, urban and rural areas present distinct exposures (17), while many biological and behavioral phenomena follow cyclical daily (18,19) or seasonal patterns (19,20). Further, some biomarkers have been demonstrated to show interaction effects between sex and other covariates (21–23). Nonlinear modeling is therefore particularly useful for capturing these complex relationships and improving the accuracy of covariate adjustment.

Here, we evaluated the impact of modeling nonlinear and interaction effects of spatiotemporal covariates on PGS predication accuracy, focusing on 16 continuous and three case-control traits available for up to 316,867 White British individuals from the UK Biobank (24). To model covariates, we adapted the null model framework from DeepNull, as it provides a modular and computationally efficient method to evaluate and integrate the contribution of non-genetic covariates. We evaluated three null model classes (linear regression, neural networks, and gradient-boosted decision trees) on different subsets of available covariates. We then integrated null models into polygenic scores using BASIL (25) and PRS-CS (26). Finally, we applied a model interpretation method to identify specific covariate interactions that explain a large fraction of phenotypic variation.

## Results

### Overview of null model evaluation framework

We began by evaluating the performance of null models, which predict a phenotype of interest based on covariates alone (**Fig. 1**). We considered three variations for each of three classes of models: linear regression, neural networks, and gradient-boosted decision trees. The linear models served as a baseline for assessing the performance improvement of the two nonlinear model classes. For this class we included a standard linear regression, as well as LASSO and ridge-penalized regressions.

**Fig. 1:**
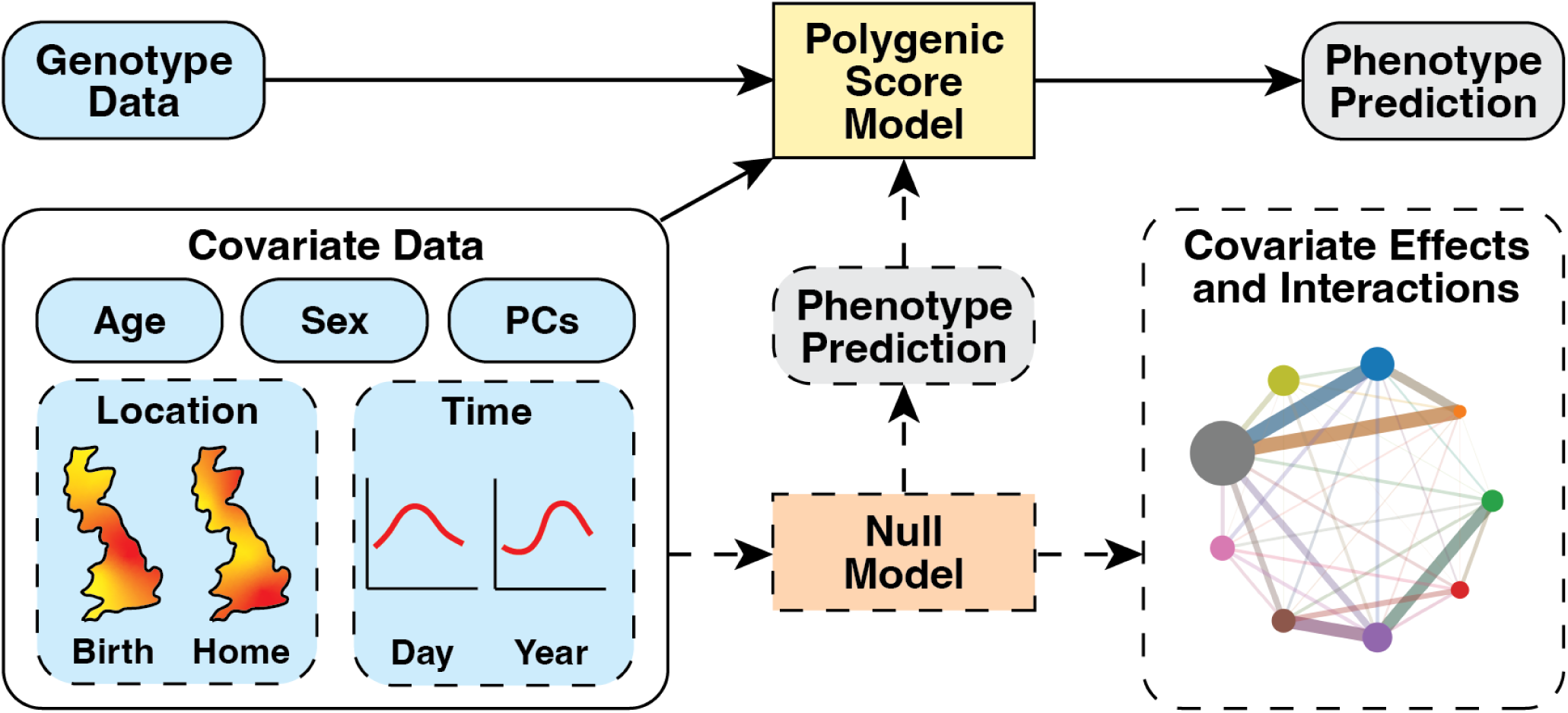
Overview of using spatiotemporal covariate null model for polygenic scores. Standard elements of a polygenic score (PGS) model are denoted with solid outlines and arrows, while additional elements considered here (spatiotemporal covariates and nonlinear null models) are indicated by dashed outlines and arrows. A standard PGS is trained to predict a phenotype based on genotype data and covariates (blue boxes). In the null model framework, we extended this by adding an auxiliary nonlinear null model (orange box), which predicts the phenotype based on the covariates alone. In addition to standard covariates of age, sex, and genotype PCs, we incorporate location and time covariates to capture spatiotemporal effects and model interpretation techniques to identify important interactions between covariates. The null model prediction (gray box with dashed border) is then used as an additional covariate input to the PGS model of choice, allowing it to capture nonlinear covariate effects and interactions.

For the neural network class, the first variation used the neural network architecture and training procedure used in the DeepNull study (5). A potential weakness of the training procedure used there is that it trained for a set number of iterations with a fixed learning rate and thus could be prone to overfitting. For the second and third neural network model variations, we therefore considered alternative training procedures that mitigate this risk by adding early stopping, weight decay, and a learning rate reduction on plateau (denoted as ESWP, see **Methods**). The second variation used the original DeepNull architecture and the third used a smaller architecture with three hidden layers instead of four.

Finally, we considered gradient-boosted decision trees, which are generally easier and cheaper to train (27,28) and which the DeepNull study reported to show similar performance to their neural networks. For this we used XGBoost (29) with early stopping. The three variations consisted of the default XGBoost parameters (a maximum of 100 estimators and a learning rate of 0.3), as well as two larger versions with up to 1,000 or 2,500 estimators, both using a learning rate reduced to 0.25 and a maximum depth of 4 to mitigate overfitting.

For each of the nine null model variations (three variations each for each class), we tested eight sets of covariate inputs. In addition to a baseline set with just age and sex, which were included in all models, the additional covariate sets added home location, birth location, time of day of sample collection, time of year of sample collection, home and birth location, time of day and year, and all the time and location covariates. We tested each of these eight covariate sets with and without the first 20 genotype principal components (PCs) included as additional covariates.

### Evaluating null model performance on continuous phenotypes in the UK Biobank

Each null model variation and covariate set was evaluated using 16 continuous phenotypes available for between 277,224-316,867 White British participants in the UK Biobank (24). These included anthropometric measurements, markers of lung, kidney, and liver function, blood pressure and lipid levels, and other blood-assay based biomarkers (**Supplementary Table 1**). We divided participants into 8:1:1 train, validation, and test sets. For each phenotype, the subset of these sets for which phenotype data was available was used (**Supplementary Table 1**). We considered a total of 2,304 models (phenotype/covariate/model variation combinations). To preserve the validation and test sets for the downstream PGS task (described below) while still providing predictions for all individuals, just the training set was used to train and evaluate the null models on their own using five-fold cross validation.

We first evaluated the performance of the different model class variations (**Supplementary Table 2**). We performed two types of evaluations. First, we considered overall performance across the different covariate sets and phenotypes. For this, we used a nonparametric ANOVA Friedman test followed by a Nemenyi post-hoc test to determine pairwise significant differences between each model class variation (30). This method is based on ranking each phenotype/covariate/model combination by performance, blocked by phenotype-covariate set pair (such that model variations were ranked separately for each of the 256 phenotype-covariate set pairs). We used R^2^ as the performance metric for each model, though the results when using mean absolute error (MAE) are similar (**Supplementary Fig. 1**).

We observed significant differences in performance across model variations (**Fig. 2a**; Friedman test P=9.1e-182). Overall, the two largest XGBoost models (maximum either 2,500 or 1,000 estimators) showed significantly better performance than any other model variations (at Nemenyi α=0.05 for all pairwise comparisons). Across all phenotype/covariate combinations, the median improvement for the largest XGBoost model compared to a linear regression null was a 4.3% increase in R^2^ and a 0.55% decrease in MAE. By average rank, the original DeepNull neural network was the second-worst performer, only outperforming the LASSO regression, and did not perform significantly differently than the linear or ridge regression models. However, the neural network models with the additional modifications to prevent overfitting performed better, with results similar to the smallest XGBoost model variation tested, which used default parameters (100 estimators) and early stopping.

**Fig. 2:**
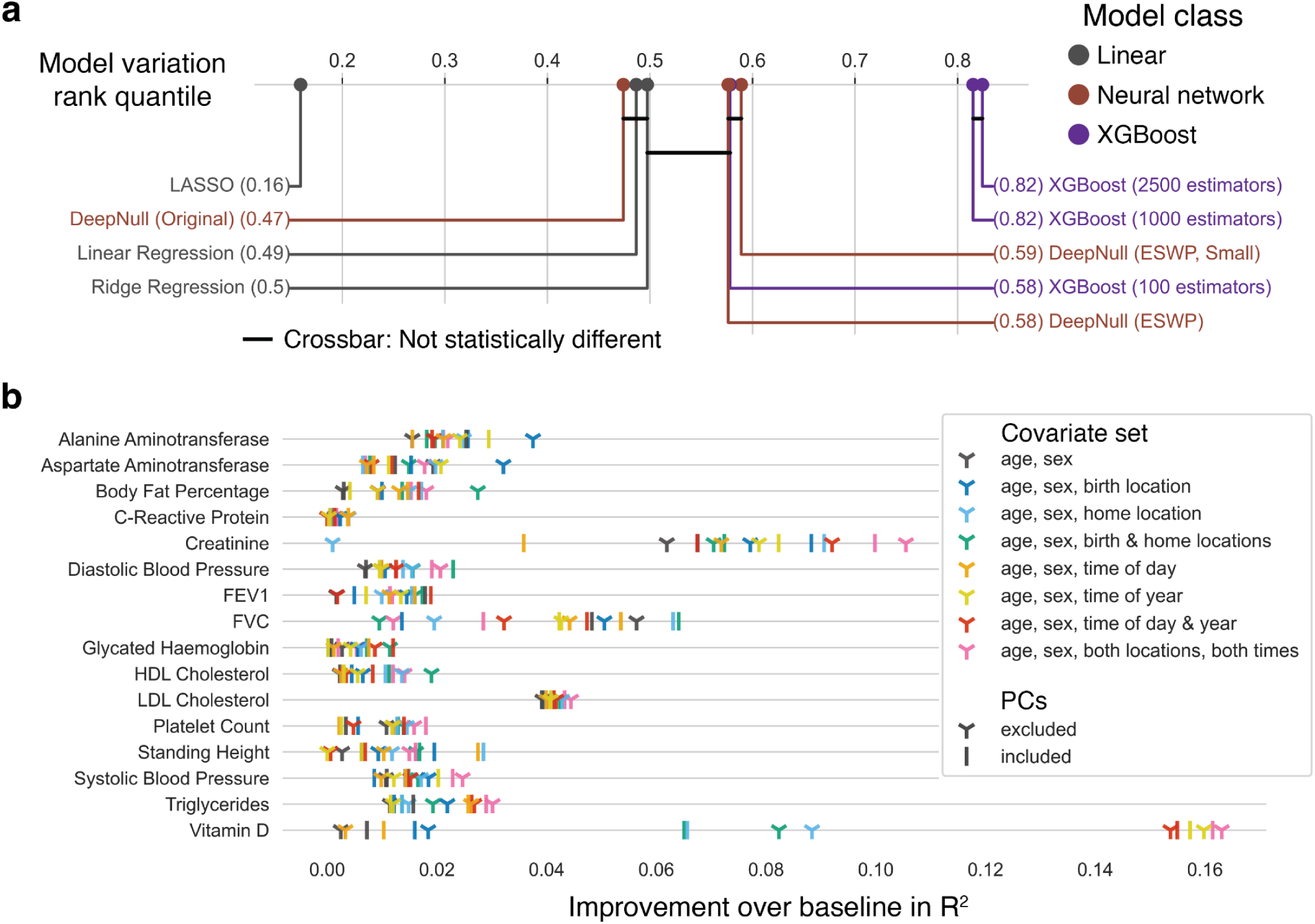
Null model evaluation results. **(a) Model variation critical difference plot.** The x-axis shows the quantile of a model variation’s R^2^ rank across all pairs, where a higher quantile indicates better predictive power on average. The horizontal crossbar connects model variates that were not statistically different with a Nemenyi post-hoc test α of 0.05 (following a significant Friedman test). The linear, neural network, and XGBoost classes of model variations are indicated with different colors. Color indicates model class (gray=linear; red=neural network; purple=XGBoost). **(b) Best null model improvement for a covariate set over the baseline approach.** The x-axis shows the improvement in R^2^ of the best performing model variation for each covariate set we evaluated compared to a baseline linear model with age and sex alone. Vertical lines indicate models with PCs included and three-pronged shapes indicate models without PCs. Color indicates covariate sets.

Second, we identified the top performing model variation for each phenotype or phenotype/covariate set pair. Overall, we observed a clear improvement in R^2^ for the nonlinear vs. linear null models. When considering the best model variation for each phenotype across all covariate sets, no phenotype had a linear model as the top performer, with a linear model only performing best for 8/256 pairs. The smaller neural network performed best most often (top model for standing height, platelet count, glycated haemoglobin, FEV1, and creatinine), followed by the larger version with the same training modifications (diastolic BP, FVC, HDL-C, and CRP). The largest XGBoost model (systolic BP, LDL-C, and triglycerides) and the original DeepNull neural network (body fat percentage, AST, and ALT) were both best for three phenotypes, and the middle sized XGBoost model was best for vitamin D.

We next compared the relative performance of different covariate sets for each phenotype, in this case blocking results by phenotype-model variation pairs to evaluate aggregate differences across all phenotypes. The covariate sets including all additional location and time features, with and without PCs, showed top overall performance. On the other hand, the baseline covariate set of age and sex was the worst performer by average rank. Adding PCs or time of day alone did not significantly outperform compared to age and sex, but most covariate sets considered showed substantial improvement (median increases in R^2^ 3.3% and 2.9% and decreases in MAE of 0.38% and 0.29%, with and without principal components respectively).

Finally, we characterized the top performing covariate set by phenotype, in each case considering the model variation/covariate-set pair with the top performance for that phenotype based on R^2^. All best performing models included additional time and/or location features. Using the additional time and location features without PCs was best for 5/16 phenotypes (vitamin D, systolic BP, LDL-C, triglycerides, creatinine). The optimal covariate set varied across other phenotypes considered, with no other set performing best for more than two phenotypes (**Fig. 2b**). Due to the variation in covariate set performance by phenotype, we continued to evaluate different phenotype set options in the following section, where we integrate the null models with the additional time and location features into PGSs.

Overall, our results demonstrate that XGBoost achieved better or similar performance compared to neural networks for this task. Additionally, the XGBoost models showed more consistent performance across the covariate set options tested (**Fig. 2a**), are less expensive and easier to train (26–28), and allow application of interpretability methods that are efficient with desirable properties (e.g. guarantees of local accuracy and consistency, generalization to any-order interactions) (7,33,34). Therefore, the following sections focus on the XGBoost model with up to 2,500 estimators as the null model but continue evaluating different covariate set options across phenotypes.

### Incorporating null models of covariates improves PGS performance

Previous work found that including the null model output as a covariate input to GWAS should improve statistical power and thus improve prediction accuracy of PGSs derived from GWAS results (5). We tested the impact of using our improved nonlinear null models for PGS construction using two separate methods. The first, PRS-CS (26), is a summary statistics-based method in which the summary statistics of a single GWAS are readjusted using an external linkage disequilibrium (LD) panel. The second, BASIL (25), is an iterative method, in which GWAS is used to select candidate variants as input to a LASSO model. This process is repeated iteratively, with the following rounds fitting to the LASSO residuals from the previous iteration. Both methods are based on linear models. For the association testing steps for each method, we incorporated the null model prediction when the overall model included a null as well as individual terms for the additional covariates described above.

We evaluated these methods with and without null models using the 16 continuous phenotypes considered in the previous section. We evaluated using the standard covariates (age, sex, and genotype PCs), in addition to location, time, and both location and time features as covariates for the PGS model. For each of these covariate options, we tested a version with and without the prediction of the null model as an additional covariate. As in DeepNull (5), covariates were included as linear terms to the PGS model even when they are already modeled by the null model since that model only captures the relationships between the covariates and the phenotype, not the covariates and the genotypes. To evaluate the relative benefits of including or excluding the PCs as input to the null model, we also tested a version using all additional covariates as well as the PCs as input. Performance for these variations for the BASIL models are shown in **Fig. 3** and for PRS-CS in **Supplementary Fig. 2**. Full results for each covariate set and model choice are provided in **Supplementary Table 3.**

**Fig. 3:**
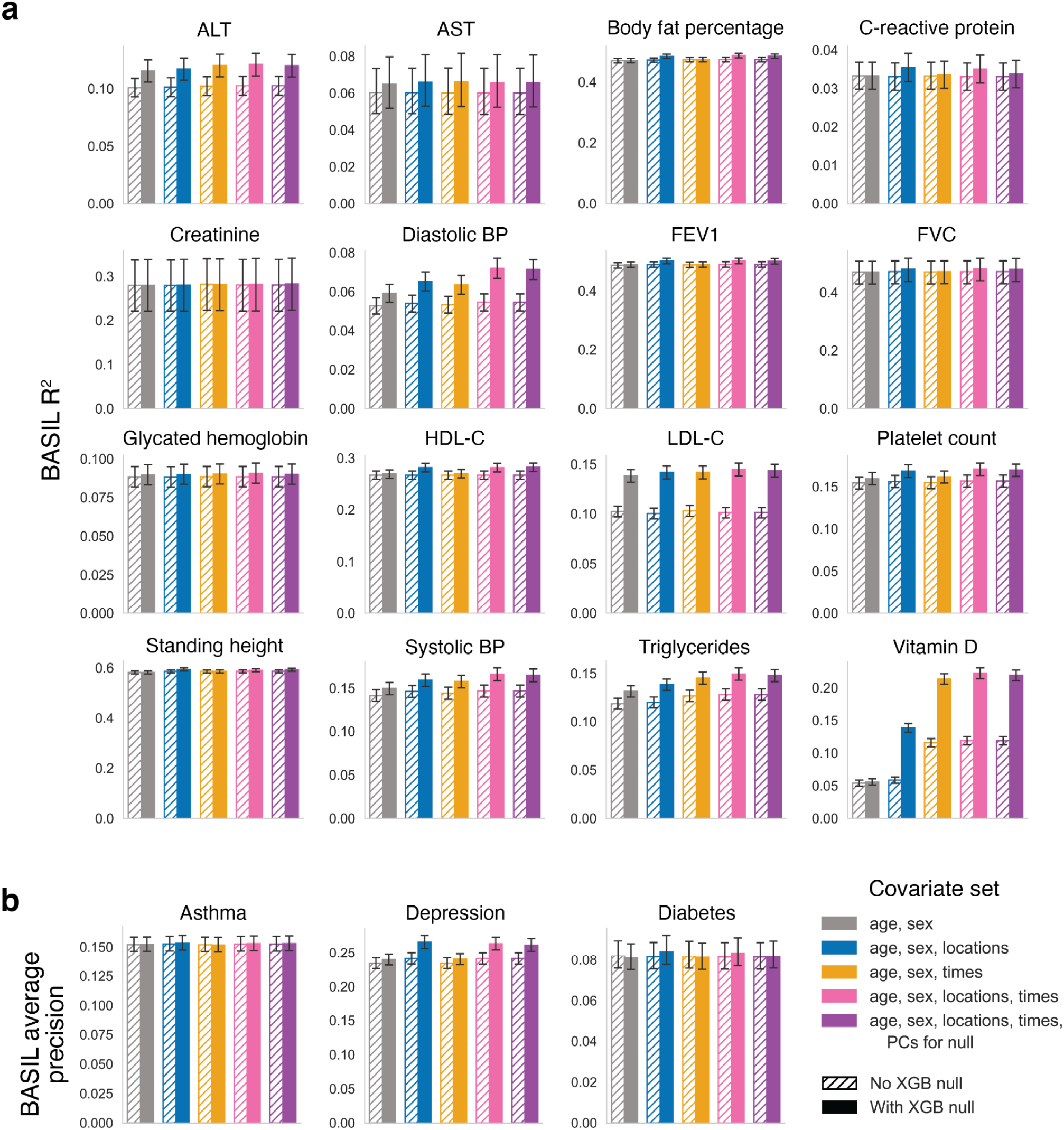
BASIL PGS model performance across model and covariate set options. **(a) Performance for continuous phenotypes.** Plots show performance of BASIL PGS models, measured by R^2^, for the 16 continuous phenotypes when the different covariate options were evaluated with and without a nonlinear null. Error bars show 95% bootstrapped confidence intervals. For each covariate set, the hatched bar shows the R^2^ when no nonlinear null was used and filled bars show performance when the nonlinear null was included. The pink and purple hashed bars are the same, as the only difference between the “age, sex, locations, times” and “age, sex, locations, times, PCs for null” covariate sets is whether the PCs are included as input to the null in addition to the other covariates. **(b) Performance for case-control phenotypes.** Plots show the performance of the BASIL PGS models for the three case-control traits. Average precision was used to evaluate performance. As for **(a)**, the error bars show 95% bootstrapped confidence intervals and the hashed pink and purple bars are the same.

For all phenotypes for both BASIL and PRS-CS, the best performance was achieved when a nonlinear null model was used with the additional time and/or location features, with the best covariate set varying across traits. For most settings tested, BASIL models outperformed those from PRS-CS, but overall trends across covariate sets and models were similar for both. For 11/16 phenotypes for BASIL and 12/16 phenotypes for PRS-CS, performance measured in R^2^ with a nonlinear null and additional spatiotemporal features was greater than the upper boundary of the 95% confidence interval of the standard approach (age, sex, and PCs as covariates without a nonlinear null model). In contrast, just adding additional covariates without a null model resulted in performance outside this confidence interval for just 2/16 traits with BASIL (vitamin D and triglycerides) and 3/16 with PRS-CS (vitamin D, triglycerides, and systolic blood pressure). There were 5 cases with BASIL and 4 with PRS-CS where a nonlinear null model with just age and sex as inputs led to performance improvements outside the 95% CI of the standard approach. Of all 128 possible cases for which we tested the same set of covariates with and without the nonlinear null model, the null model only showed worse performance in two cases, both of which had R^2^ decreases of less than 0.0005. Overall, these results show that incorporating a null model with spatiotemporal features in almost all cases improves or does not harm performance. Further, the combination of spatiotemporal covariates with nonlinear null models provided greater improvement over incorporating either these additional covariates or null models alone.

The covariate set that showed top performance (best R^2^) for the majority of phenotypes was including the nonlinear null model with the additional location and time covariates (top for 11/16 phenotypes with BASIL and 12/16 phenotypes with PRS-CS). For BASIL and PRS-CS respectively, the median improvement in R^2^ for this approach was 7.3% and 15.5% compared to the standard approach of just including age, sex, and PCs as covariates to the PGS model, 7.5% and 11.4% compared to additionally including the location and time covariates as linear covariates to the PGS model, and 4.7% and 6.2% compared to using age and sex as covariates with a nonlinear null model. The optimal covariate set varied across phenotypes. For example, for BASIL there were two phenotypes (C-reactive protein and standing height) for which the best performance was achieved when location but not time features were included, and one (aspartate aminotransferase) for which the best performance was when time but not location was included. BASIL PGS models with nonlinear nulls achieved R^2^ outside the 95% CI of the standard approach for 11 of 16 continuous phenotypes each when using either additional location, location and time, or location, time, and PC features. For PRS-CS, this was for 12, 11, and 11 phenotypes, respectively.

We also evaluated how the nonlinear null models and additional spatiotemporal covariates impacted the performance of BASIL for three case-control traits from UK Biobank: asthma, depression, and diabetes. For these phenotypes we used 254,008 labeled samples with case control ratios of 1:6.05, 1:4.59, and 1:18.2 respectively. For all three phenotypes, the best results occurred when a null model was used with the addition of location features, though the degree of impact varied. Depression was the only phenotype for which there was improvement in average precision and area under the receiver operating curve (auROC) outside of the 95% confidence interval of the standard approach. The average precision improved 13.5% over the standard approach and 10.1% compared to including the location features but only modeling them linearly. The improvements in auROC were 6.1% and 5.4% respectively. For asthma and diabetes, the improvements in average precision and auROC were minor and never outside of the 95% confidence interval of the standard approach.

Ultimately, for continuous traits we found that adding a nonlinear null and additional spatiotemporal features to PGS models were both advantageous, but using both together performed best. Using both resulted in performance outside the 95% confidence interval of the baseline for more than twice as many phenotypes compared to applying either of those strategies individually. The improvements in terms of R^2^ achieved by null models with spatiotemporal features over baseline models is highly correlated with the improvement in R^2^ for BASIL PGSs when the nonlinear null and spatiotemporal features are added (Pearson correlation = 0.9985, p-value 5.4e-19) (**Supplementary Fig. 3**). For the three case-control phenotypes we did not see a clear trend of improvement, but the one example for which a model exceeded the 95% confidence interval of the baseline was when location features and a nonlinear null were used in conjunction.

### Identifying covariate interactions with strong contributions to null models

Unlike linear models, nonlinear models can be used to understand interactions between their input features in addition to the inputs’ separate additive effects. Given the notable improvements in null model and PGS performance using models that account for nonlinear effects and covariate interactions, we next sought to identify specific interactions within our extended set of covariates that are important for explaining phenotypic variation. Here we focused on XGBoost, which we used above in PGS models. These GBDT models are a good fit for understanding both effects of individual covariates and interactions, since interpretability methods exist that can efficiently compute effects of individual factors, first-order, and higher-order interactions by taking advantage of the underlying tree-based structure of the model (7,34)

We used TreeSHAP-IQ (34) to compute first-order Shapley Interaction Index (1-SII) values (35) to identify covariate interactions. One complication when using Shapley value based interpretability methods is that they provide local (i.e. for one individual) measures of impact or importance when we are mostly interested in understanding the global importance, which reflects the aggregate importance across the population. To identify covariate interactions that were globally important, we looked at the mean magnitude of the local first-order SII values for the held out test set. We then visualized these global relationships using network plots similar to those used by TreeSHAP-IQ for visualizing local feature attribution in addition to examining 1-SII values for individual features and pairs of features (**Fig. 4; Supplementary Fig. 4**).

**Fig. 4:**
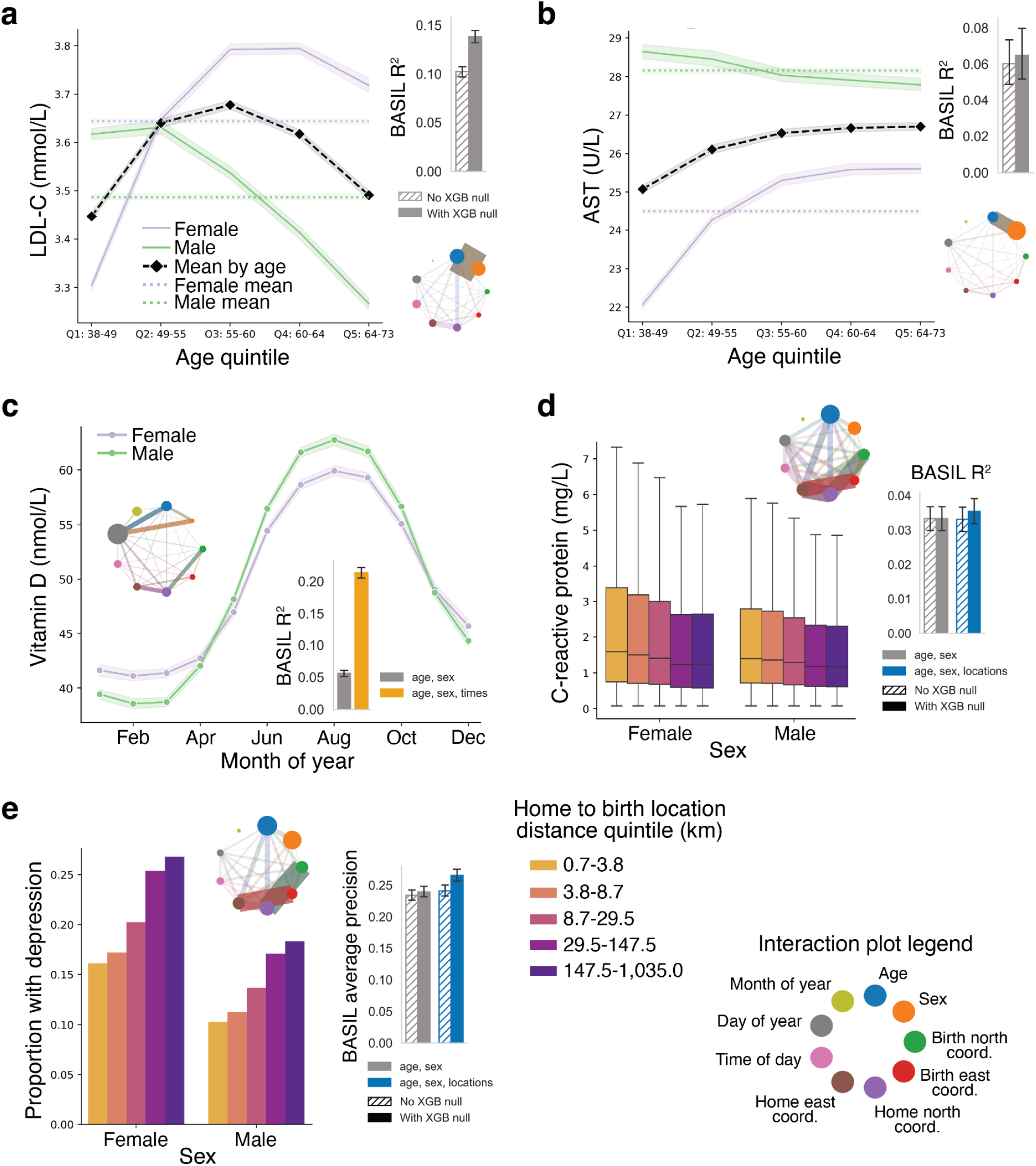
Covariate interactions identified by null model interpretation. **(a) Age-sex interactions for LDL cholesterol (LDL-C).** The main plot shows mean LDL-C levels for males and females in each of the five age quintiles, in addition to the overall mean for males, females, and each age group separately. Shading indicates 99% confidence intervals. The bar plot (top right) shows the BASIL PGS model’s improvement in performance when a nonlinear null was used, thus allowing it to model these interactions. The bottom right panel shows a covariate interaction network plot defined below. **(b) Age-sex interactions for aspartate aminotransferase (AST).** Panels are the same as in **(a)** except for AST. **(c) Sex-time of year interaction for vitamin D.** The main plot shows the variation in average vitamin D levels for males and females over the course of the year. Note that this plot shows the levels binned by month instead of day of year, which is collinear but identified as having a greater impact by TreeSHAP-IQ, as seen in the interaction plot in the upper left. Shading shows 99% confidence intervals. The bar plot shows the PGS improvement when the time covariates are included in addition to age and sex when a nonlinear null is used, which can capture this cyclical nonlinear behavior. **(d) Birth-to-home distance interaction for C-reactive protein (CRP).** The main box plot shows the CRP levels for each of the five birth-to-home distance quintiles for males and females. The legend below **(d)** shows the colors corresponding to the birth-to-home distance quintiles used in **(d)** and **(e)**. The whiskers extend to the farthest datapoint within 1.5 times the interquartile range (IQR) from the boundary of the middle two quartiles. The interaction network plot is shown above the box plot. The bar plot showing the improvement when location features are modeled with and without a nonlinear null is to the right. **(e) Birth-to-home distance interaction for depression.** The bar plot shows the proportion of males or females in a birth-to-home distance quintile that reported past or current depression. The interaction network plot is provided along with a bar plot of BASIL polygenic score performance, which shows the largest improvement when location features are included and modeled nonlinearly. For all bar plots, error bars indicate 95% bootstrapped confidence intervals. Hatched and solid bars denote models without and with null models, respectively. For interaction plots, circle sizes indicate the mean magnitude of SHAP values for each covariate and edge widths indicate the mean magnitudes of 1-SII values for pairwise covariate interactions.. All 1-SII values for individual covariates and pairwise interactions for all traits are shown in **Supplementary** Fig. 4.

We identified several influential categories of interactions contributing to the traits studied. First, we found multiple traits (ALT, AST, LDL-C, triglycerides, vitamin-D) where the age-sex interaction had a greater mean 1-SII magnitude than age and/or sex alone (**Fig. 4a-b**; **Supplementary Fig. 4a, b, n, r, s**). For some phenotypes (ALT, AST, LDL-C, and triglycerides), the trajectories with age are largely in opposite directions for males vs. females. These interactions are similar to those shown by studies in other cohorts for LDL-C and triglyceride levels (21), ALT (36), and systolic blood pressure (37). LDL-C (**Fig. 4a; Supplementary Fig. 4n**) most clearly demonstrates the importance of modeling age and sex covariates in a way that accounts for age-by-sex interactions. If sex alone were considered, females would be deemed to have higher LDL-C levels. On the other hand, when considering age, it is clear that females’ levels are only higher on average in and after the second age quintile (ages 49-55), likely as a result of menopause (21). If age alone were considered, the different trajectories for males and females with age would be missed. The importance of this interaction is reflected in the mean 1-SII magnitudes for LDL-C, where the value for the age-sex interaction was greater than age or sex alone (**Supplementary Fig. 4n**). Aspartate Aminotransferase (AST) provides a similar example: considering age alone would miss the opposite trend observed for males compared to females (**Fig. 4b**). The mean 1-SII magnitude for the age-sex interaction is less than that of sex but greater than age (**Supplementary Fig. 4b**).

Second, we identified a strong interaction between sex and time of year for vitamin D levels (**Fig. 4c**). The sex-day of year interaction had the sixth highest mean 1-SII magnitude (**Supplementary Fig. 4s**). Vitamin D levels show a sinusoidal pattern, with peak values in August and lowest levels in February. The means for males and females are similar (less than a tenth of a nanomole per liter difference), but the amplitude of the sinusoidal pattern for males is larger. This is supported by other studies that report greater vitamin D variability over the course of the year for men than women, possibly explained by lower levels of adipose tissue (15). This time-dependent variability may also explain why other studies looking at sex differences in vitamin D levels without accounting for time of year are inconsistent (15).

Finally, we identified multiple strong interactions involving location. For some traits, we observed interactions between home and birth location coordinates. This indicated there may be some relationship between where someone lives compared to where they were born and the phenotype. For example, for C-reactive protein (CRP) the birth and home east coordinate interaction ranked fifth highest in mean 1-SII magnitude and the birth and home north coordinate interaction ranked seventh (**Supplementary Fig. 4e**). Looking into this further, we found the birth-to-home distance is negatively correlated with CRP levels (Spearman r = -0.077; P<10^-100^). This trend is seen for both males and females when visualizing CRP distributions by birth-to-home location quintile (0.7-3.8 km, 3.8-8.7 km, 8.7-29.5 km, 29.5-147.5 km, and 147.5-1,035.0 km from their birth location; **Fig. 4d**).

In another example, for current or prior depression the self-reported rate increased with increased birth-to-home location distance (**Fig. 4e**). This result matched the findings of a 2023 study on internal migration in the United States, which found that the rate of depression among internal migrants was 1.259 times higher than for nonmigrants (38). We found significant positive correlations between depression and birth-to-home location distance (point-biserial r = 0.076, rank-biserial r = 0.15, both P-values < 10^-100^). The birth and home north coordinate interaction had the third highest mean 1-SII magnitude following only age and sex on their own (**Supplementary Fig. 4g**). The birth and home east interaction was fifth highest, one behind the home north coordinate. Importantly, our results do not distinguish if the cause is due to differences in attributes between those who do or do not migrate or properties of the locations involved. Still, our results help explain why incorporating interactions between features can improve PGS prediction accuracy.

## Discussion

In this study, we demonstrate that improved modeling of the non-genetic component of traits can substantially enhance the performance of PGSs. Our results highlight that the combination of spatiotemporal features and nonlinear null models leads to more significant improvements in PGS accuracy than either modification on its own. We found this to be particularly true for the continuous phenotypes tested here, where the combination of these two improvements resulted in the best performance for all phenotypes and R^2^ outside of the 95% confidence interval of the baseline for 11/16 traits with BASIL and 12/16 with PRS-CS. For the three case-control phenotypes we evaluated we saw modest differences in performance, except for depression where the combination of a nonlinear null model and location features resulted in performance outside the baseline 95% CI.

The utility of specific spatiotemporal covariates was phenotype-dependent. For phenotypes known to have diurnal or seasonal rhythms, like vitamin D or blood pressure, the time-based covariates were highly informative. Conversely, for standing height, which is highly heritable and for which impact of environmental exposures likely occur before adulthood (39), as expected adding factors such as time of assessment have no impact on performance. Similarly, for the case-control phenotypes, the time of assessment was not relevant to whether a person would have previously developed any of the conditions, and as expected they had either no effect or a detrimental effect on performance. In other case-control applications like longitudinal studies, time features could be more relevant for understanding case-control traits. In practice, researchers should consider the underlying dynamics of a trait when selecting covariates or compare the performance null models, which are relatively cheap and easy to train, to evaluate different covariate sets. Nonetheless, for the majority of continuous traits we tested, a model incorporating the full set of spatiotemporal covariates performed best.

A strength of the null model approach is that, beyond improving prediction performance, it provides insight into how non-genetic factors and their interplay impact different traits. By modeling nonlinear and interaction effects among covariates, these models reveal dependencies that linear adjustments fail to capture. In this study, we detected both novel and previously reported covariate interactions, including examples where phenotype means exhibit opposite age-related trends between sexes (ALT, AST, LDL-C, and triglycerides), cyclical temporal patterns for vitamin D reflecting seasonal variation, and complex spatial dependencies between birth and home locations. These location differences can capture effects of distance or direction moved, as well as nonadditive interactions between the effects of where one grew up and now lives. Together, these findings illustrate how nonlinear null models can uncover interpretable relationships that are obscured under standard linear frameworks.

Our study has several limitations. First, the analysis was conducted exclusively on individuals of White British ancestry in the UK Biobank. The specific environmental patterns learned by our models are likely population-and location-specific and may not generalize to other ancestral groups or geographical regions. Second, spatiotemporal variables are in most cases proxies for true environmental exposures. Our models and methods for interpreting them cannot determine what the specific causal factor is. For example, if location is determined to be important, we still would not know if that is due to the weather, pollution, or lifestyle differences in an area. Third, while our method improved predictions for continuous traits, the benefit for case-control traits was modest. This could be due to a smaller environmental contribution for the selected phenotypes, lower statistical power, or challenges in phenotype definition (e.g., self-reports of past instances of depression).

In our study we focused on integrating null models into commonly used classes of PGSs based on linear models. In this approach, the null models are trained separately on covariates, and the resulting output is used as an additional linear term in association testing or PGS construction. As discussed in the introduction, an alternative approach is to perform feature selection on the genotypes and then use a nonlinear model to predict the phenotype based on a model that jointly considers the subset of genotypes and covariates (12–14). In this setup the feature selection is a key step that could be similarly improved with the integration of a null model of covariates, which is an important topic of future work.

Overall, we demonstrate that incorporation of nonlinear and interaction effects of spatiotemporal covariates into association testing models can in some cases result in large performance gain with relatively small additional computational overhead beyond standard approaches. Importantly, our framework, which builds on DeepNull (5), outputs a single term summarizing these covariate contributions that can be easily added as a covariate to any downstream methods for complex trait analysis, such as genome-wide association studies, PGS, and fine-mapping. Further, location and time related covariates are already collected by most large biobanks and the gradient boosted decision tree null models are relatively cheap and easy to train. This makes our approach easy to implement for future studies. Ultimately, this work demonstrates the importance of modeling the environmental component of complex traits and provides a practical framework for integrating these features into large-scale genetic studies to improve prediction and discovery.

## Methods

### Participant data

All analyses were performed using data from the UK Biobank (UKB) (24) on the UKB Research Analyses Platform. We excluded participants who did not self-identify as ’White British’ (field 22006), did not meet the quality control (QC) criteria for inclusion in the UKB principal components computation (field 22020), were labeled as outliers for heterozygosity or missing rate (field 22027), had sex chromosome aneuploidy (field 22019), or exhibited sex discordance (fields 31 and 22001). This resulted in 337,081 participants. These individuals were used for the genotype QC, during which additional participants were also removed based on their genotype missingness rate (see below). To avoid having missing covariate values, we further subsetted to the 317,511 participants for which home and birth location data were available for all analyses. These remaining participants were split into 8:1:1 training, validation, and test sets. The final sample counts per phenotype are provided in **Supplementary Table 1.**

### Genotype data

We used the version 3 UKB imputed genotypes. The following genotype QC steps were performed with plink2 v2.00a2.3LM (40,41) using the sample set described above: We began by removing variants with a minor allele frequency less than 0.01, an INFO score of less than 0.3, or that were multiallelic. A coarse missingness filter was then applied, removing participants and then variants with a missingness rate greater than 20%. Finally, variants with a missingness rate greater than 2% were removed, followed by participants with a missingness rate greater than 2%.

### Covariate data

Covariate information was derived from UKB fields. Following the sample QC, genotype QC, and removal of samples with missing location data, there were no missing values. The age covariate was obtained from the age at recruitment (field 21022), which reflects a participant’s age at their initial assessment center visit. Sex was coded as a binary variable (field 31). We used the first 20 genetic principal components, as provided by UKB in field 22009.

For location covariates, we considered participants’ home and birth locations. These features are encoded as easting-northing coordinate pairs using the Ordnance Survey National Grid (OSGB) reference system (fields 130 and 129 for birth, and 22702 and 22704 for home).

For time covariates, we included both the time of day and the time of year based on the time the blood sample was collected during the initial assessment center visit (field 3166). Time of day was represented continuously in terms of minutes past midnight. Time of year was represented with integers for both month and day of the year.

### Phenotype data

Sixteen continuous phenotypes were used, each derived from the initial assessment visit for a single UK Biobank field: alanine aminotransferase (field 30620), aspartate aminotransferase (field 30650), body fat percentage (field 23099), C-reactive protein (field 30710), creatinine (field 30700), diastolic blood pressure (field 4079), forced expiratory volume in 1-second (FEV1, field 3063), forced vital capacity (FVC, field 3062), glycated haemoglobin (field 30750), HDL cholesterol (field 30760), LDL direct (LDL-C, field 30780), platelet count (field 30080), standing height (field 50), systolic blood pressure (field 4080), triglycerides (field 30870), and vitamin D (field 30890).

Three case-control phenotypes were used. Asthma cases were defined using the version 2.0 algorithmically defined outcomes (ADOs) made available by UKB. Participants were classified as cases if they had a value for “source of asthma report” (field 42015), which included self-reported only, hospital primary, death primary, hospital secondary, or death contributory. Depression cases were defined based on responses to the online mental wellbeing questionnaire. Participants were classified as cases if they had a value for “duration of worst depression” (field 20438). Values in this field indicated participants had answered “yes” to either “Have you ever had a time in your life when you felt sad, blue, or depressed for two weeks or more in a row?” (field 20446) or “Have you ever had a time in your life lasting two weeks or more when you lost interest in most things like hobbies, work, or activities that usually give you pleasure?” (field 20441). Diabetes cases were identified using self-reported diagnosis of diabetes by a doctor (field 2443, “yes” for any instance).

### Null models

The null models, which predict a phenotype based solely on covariates, were trained using the training set of participants with 5-fold cross-validation, as in DeepNull (5). Predictions for training set participants were from the fold in which they were held out. Predictions for members of the validation and test sets were computed as the average of the predicted values across the five folds. When evaluating null model performance on its own, we used only the held-out predictions for the training set to avoid biasing results when integrating the null model into PGSs (below).

We evaluated three classes of null models: linear, XGBoost, and neural network. The linear models were implemented using scikit-learn (42). The linear regression used the default parameters. The LASSO and ridge regression models used the default parameters except for α, which was set to 0.5 and 0.1 respectively.

XGBoost (29) is an implementation of gradient-boosted decision trees (GBDTs). We used the Python API default parameters except where otherwise noted. For all model variations, 10% of the participants in the folds used for training were held out as a validation set for early stopping. Root mean square error (RMSE) was used as the early stopping evaluation metric. The smallest of the three variations we evaluated used up to 100 boosting rounds (i.e. estimators) and stopped early if there was no improvement in 25 rounds. The two larger versions used up to 1,000 or 2,500 boosting rounds and waited 50 rounds before early stopping. To further avoid overfitting, the two larger versions used a maximum tree depth of 4 and reduced the learning rate from 0.3 to 0.25.

The neural network models were implemented using PyTorch (43). One variation was a reimplementation of the original DeepNull (5) network architecture and training procedure. The architecture has two components: a linear residual connection and a multi-layer perceptron (MLP) with four hidden layers of sizes 64, 64, 32, and 16 with Rectified Linear Unit (ReLU) activations between them. Both components take the input variables as input and their outputs are summed to get the output. The Adam optimizer (44) was used with β_1_ = 0.9 and β_2_ = 0.99. Training was run for 1,000 epochs with a batch size of 1,024 and a learning rate of 10^-4^. Mean squared error (MSE) loss was used for regressions and binary cross entropy for classification.

The other two variations modified the training procedure to help prevent overfitting. Both used early stopping, in which 10% of the participants in the folds used for training were held out as a validation set. These variations trained for up to 5,000 epochs but stopped early and reverted to their best-performing weights if the loss did not improve for 75 epochs. The AdamW optimization algorithm (45) was used with the default PyTorch parameters due to its weight decay regularization. Training was run with a batch size of 1,024 and an initial learning rate of 10^-4^. The learning rate was reduced by a factor of 0.1 if training plateaued (the loss did not improve by more than 10^-4^ for more than 25 epochs). These versions are indicated as ESWP in **Fig. 2**, referring to early stopping, AdamW, and plateau. One variation used the same architecture as the original DeepNull model while the other instead used three hidden layers of sizes 64, 32, and 16.

### Polygenic score models

The PGS models, which used either BASIL (no version information available) (25) or PRS-CS v1.1.0 (26), predicted a phenotype based on genotypes, genetic principal components, non-genetic covariates, and in some cases the prediction of a nonlinear null model. Evaluation was performed using the test set. Before these models were fit, the location coordinates, time of day, day of year, and null model predictions were standardized to have a mean of zero and a standard deviation of one. For PRS-CS, month of year was treated as a categorical variable, while for BASIL it was left as an integer. This was done as PRS-CS can accept categorical input variables.

BASIL models were trained on the training set of participants, while the validation set was used for early stopping and selecting the λ hyperparameter used in BASIL. Training was performed for up to 25 iterations, with early stopping if the validation metric did not improve for more than two iterations. An elastic-net mixing parameter of α=1.0 was used, corresponding to the LASSO penalty. The default parameters of the authors’ implementation were otherwise used.

PRS-CS is a summary statistics based method, so we began by running an association test with plink2 v2.00a6LM (40,41) using the generalized linear model (--glm flag) on the training set. The only departure from the default parameters was increasing the maximum variance inflation factor to 10^6^. This change allowed us to run the association test despite the correlation between covariates, especially the null model prediction and the other covariates it is based on, being high. The genotypes and covariates, optionally including the null model prediction, were used as inputs to the association test, though the following PRS-CS step only had the summary statistics for the genotypes as input. For PRS-CS, the 1,000 Genomes European LD reference panel provided on the PRS-CS website was used. This was selected over the UKB LD reference panel since it was orthogonal to the samples in our analysis. Participants were then scored using the posterior effect sizes from PRS-CS. To reincorporate the covariates and rescale the score to the actual phenotype, a linear wrapper model was then fit using the validation set. This linear model used either a scikit-learn (42) linear regression for continuous traits or a logistic regression for case-control traits with the default parameters. The input variables were the score from summing the PRS-CS effects, the covariates, and the optional null model prediction. The covariates were scaled as described above, except that months were instead one-hot encoded.

### Null model interpretation

We used TreeSHAP-IQ (34) to interpret the trained XGBoost null models to identify covariate effects and interactions. This was done using the test set of participants. For each participant, this resulted in SHAP values for each covariate and a matrix of first-order Shapley Interaction Index (1-SII) values. This matrix provides the 1-SII value for each covariate on its own as well as every pairwise interaction. Both the SHAP and 1-SII values are local, meaning they reflect the impact of a covariate for one participant. To make these values global (i.e. reflect their impact across the entire cohort), we used the average magnitude of the 1-SII values when identifying important interactions. Raw 1-SII values for all traits are provided in **Supplementary Fig. 4**.

## Acknowledgements

This research was supported by NIH grant RM1HG011558 (M.G.) and used the UK Biobank Resource under application number 46122 and data provided by patients and collected by the NHS as part of their care and support. We gratefully acknowledge UK Biobank participants for their contributions, without whom this research would not have been possible. We thank Tara Mirmira and Michael Lamkin for helpful comments on the manuscript.

## Code availability

Code used for this study is available at https://github.com/RossDeVito/DeeperNull.

## Data availability

We used genetic and phenotype datasets provided by UK Biobank under application number 46122 (see https://www.ukbiobank.ac.uk/use-our-data/apply-for-access).

## Author contributions

R.D. and M.G. conceptualized the project. R.D. performed all analyses and experiments, prepared the figures and wrote the manuscript. M.G. supervised analyses and helped with writing the manuscript. Both authors reviewed and approved the final manuscript.

