## Supplementary Figures for "Modeling nonlinear and interaction effects of spatiotemporal and other non-genetic factors improves phenotypic prediction for complex traits"

#### Supplementary Figure 1

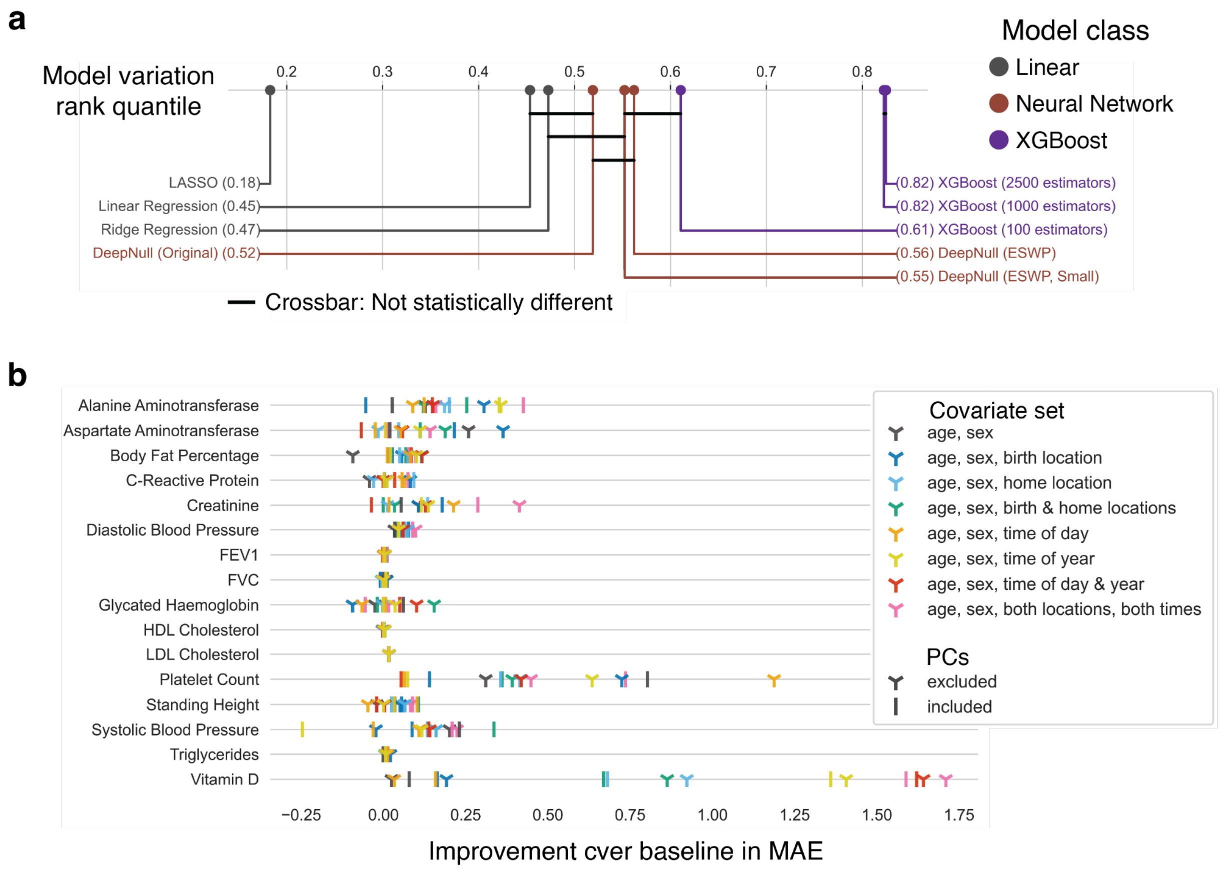


**Supplementary Figure 1: Null model evaluation results using mean absolute error (MAE).** **(a) Model variation critical difference plot.** The x-axis shows the quantile of a model variation's MAE rank across all pairs, where a higher quantile indicates better predictive power on average. The horizontal crossbar connects model variates that were not statistically different with a Nemenyi post-hoc test α of 0.05 (following a significant Friedman test). The linear, neural network, and XGBoost classes of model variations are indicated with different colors. Color indicates model class (gray=linear; red=neural network; purple=XGBoost).
**(b) Best null model improvement for a covariate set over the baseline approach.** The x-axis shows the improvement in MAE of the best performing model variation for each covariate set we evaluated compared to a baseline linear model with age and sex alone. Vertical lines indicate models with PCs included and three-pronged shapes indicate models without PCs. Color indicates covariate sets.

#### Supplementary Figure 2

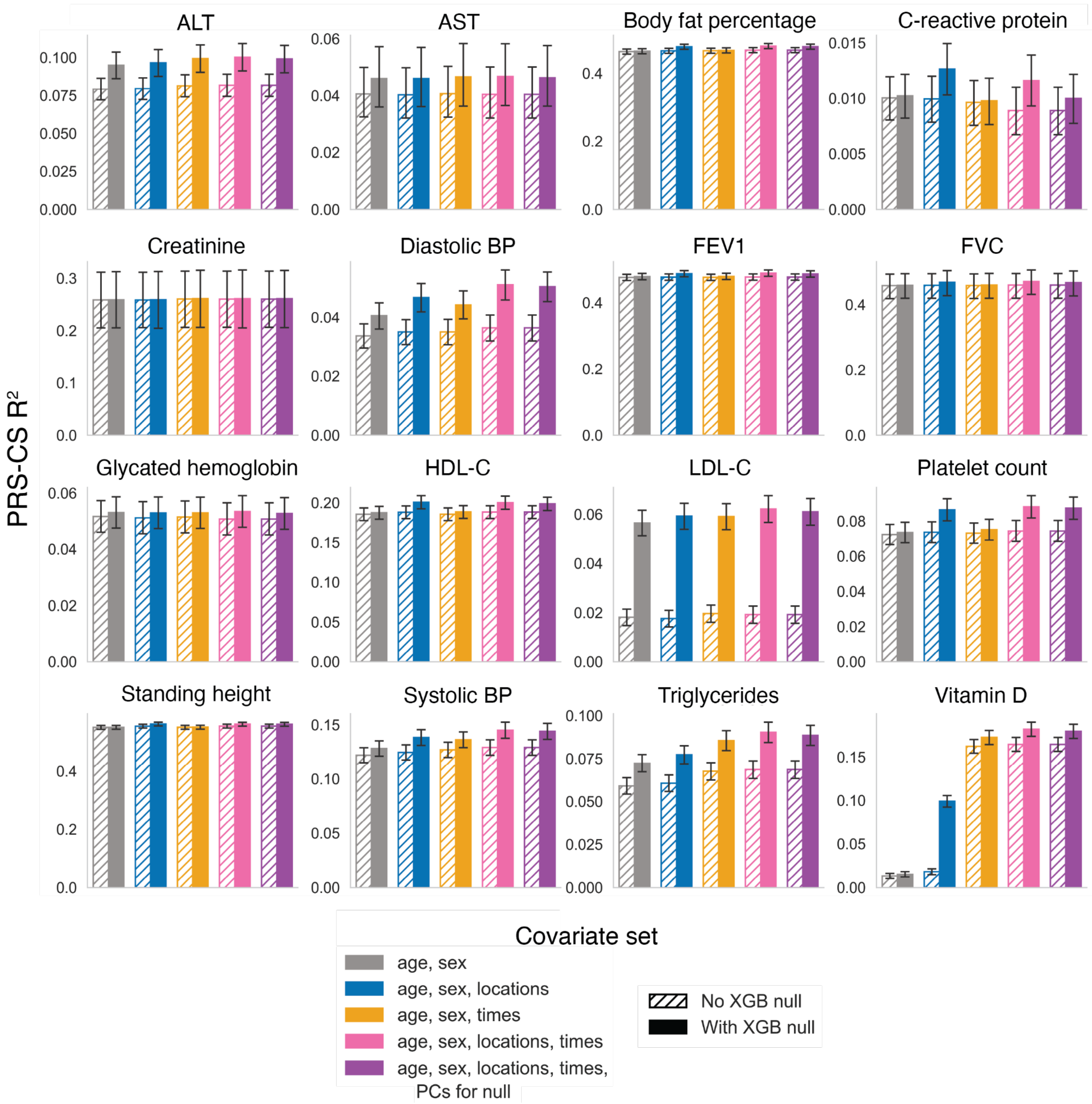


**Supplementary Fig. 2: PRS-CS PGS model performance for continuous phenotypes.** Plots show performance of the PRS-CS polygenic score models, measured in $R^2$, for the 16 continuous phenotypes when the different covariate options were evaluated with and without a nonlinear null. Error bars show 95% bootstrapped confidence intervals. For each covariate set, the hatched bar shows the $R^2$ when no nonlinear null was used and filled bars show performance when the nonlinear null was included. As in **Fig. 2**, the pink and purple hashed bars are the same, as the only difference between the "age, sex, locations, times" and "age, sex, locations, times, PCs for null" covariate sets is whether the PCs are included as input to the null in addition to the other covariates.

#### Supplementary Figure 3

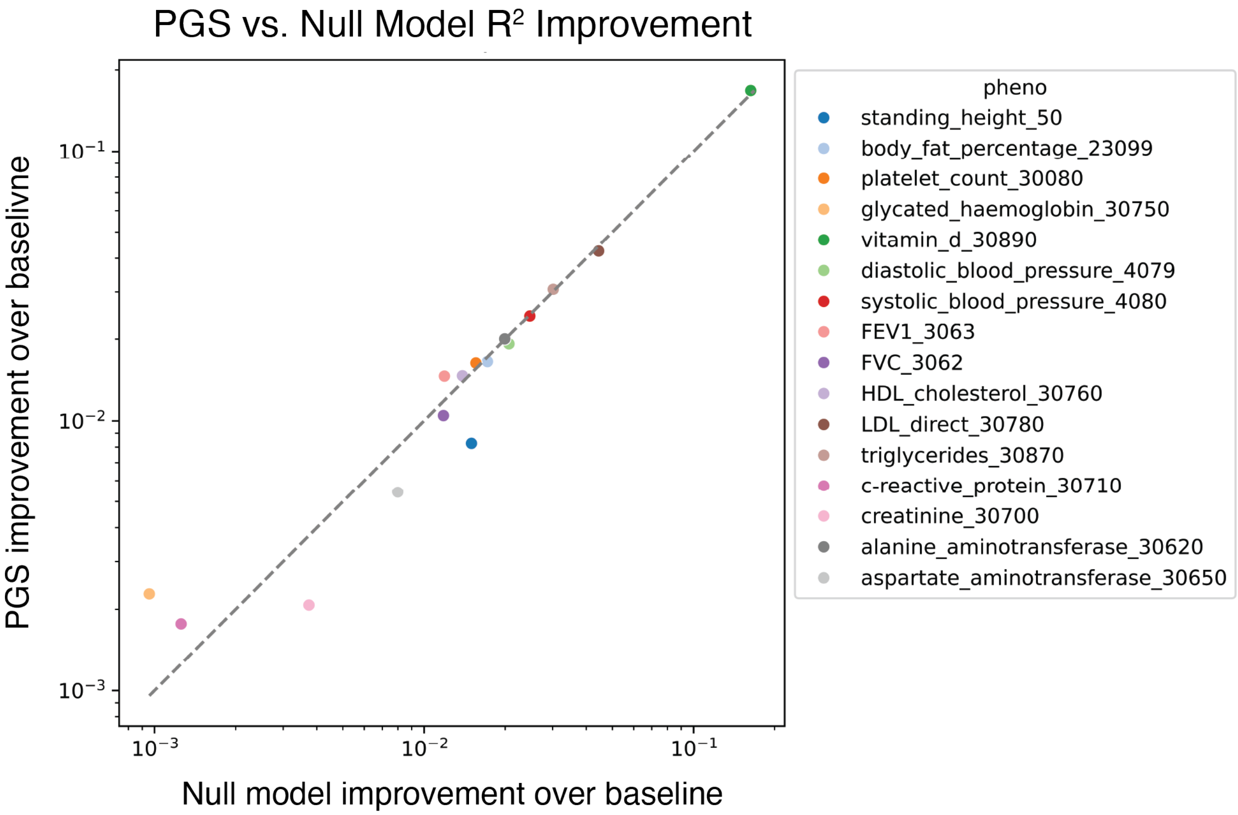


**Supplementary Fig. 3: Correlation between null and PGS improvements over baseline with spatiotemporal covariates and nonlinear null models.** The x-axis shows the improvement in $R^2$ of the largest XGBoost nonlinear null model using all the additional spatiotemporal covariates over the baseline approach (a linear regression null model using just age and sex). The y-axis shows the improvement in $R^2$ of a BASIL PGS that uses the nonlinear null and additional spatiotemporal features over one that does not. The Pearson correlation of these improvements is 0.9985 with a p-value of 5.4e-19.

### Supplementary Figure 4

#### Supplementary Figure 4a

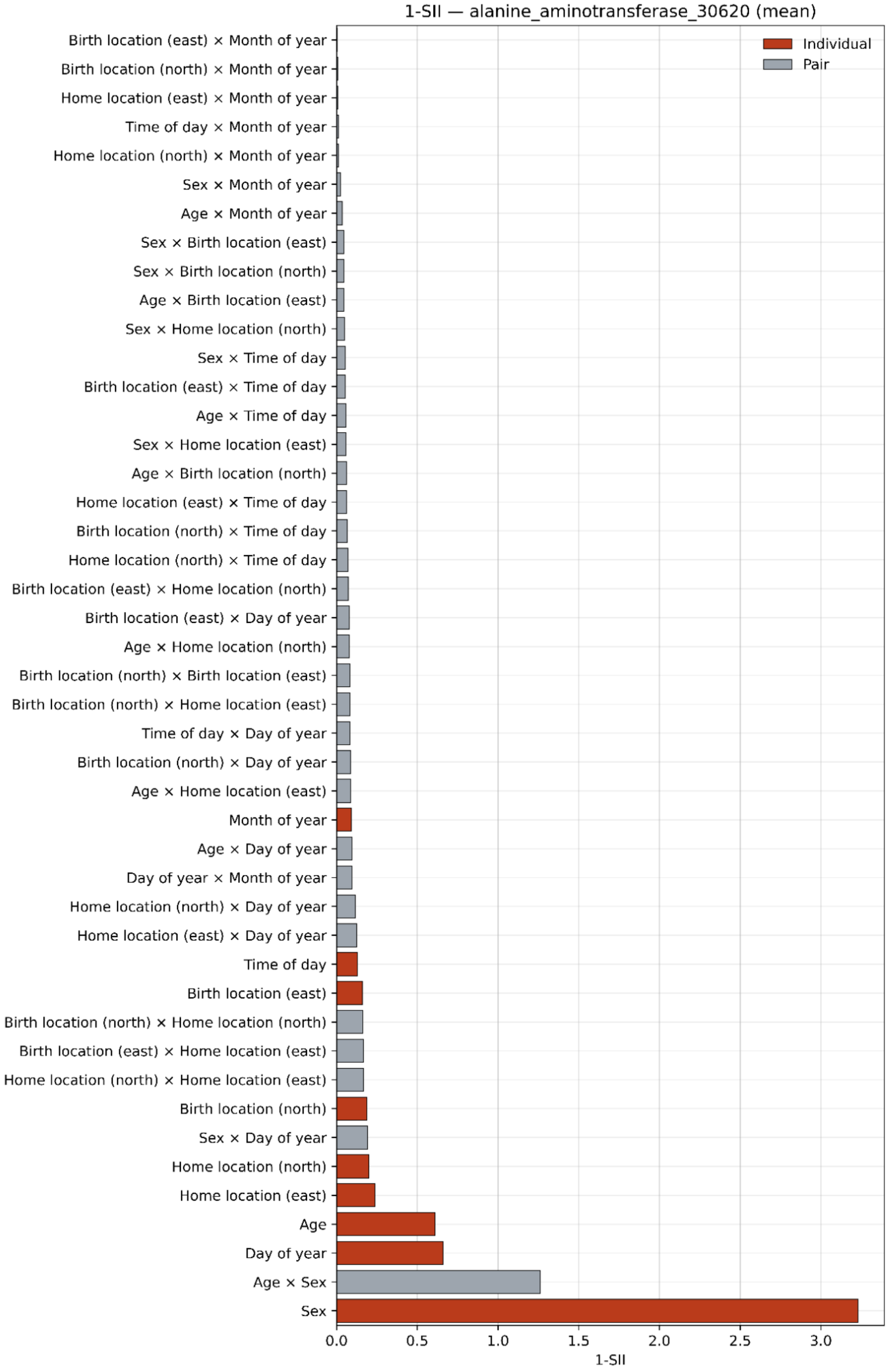

#### Supplementary Figure 4b

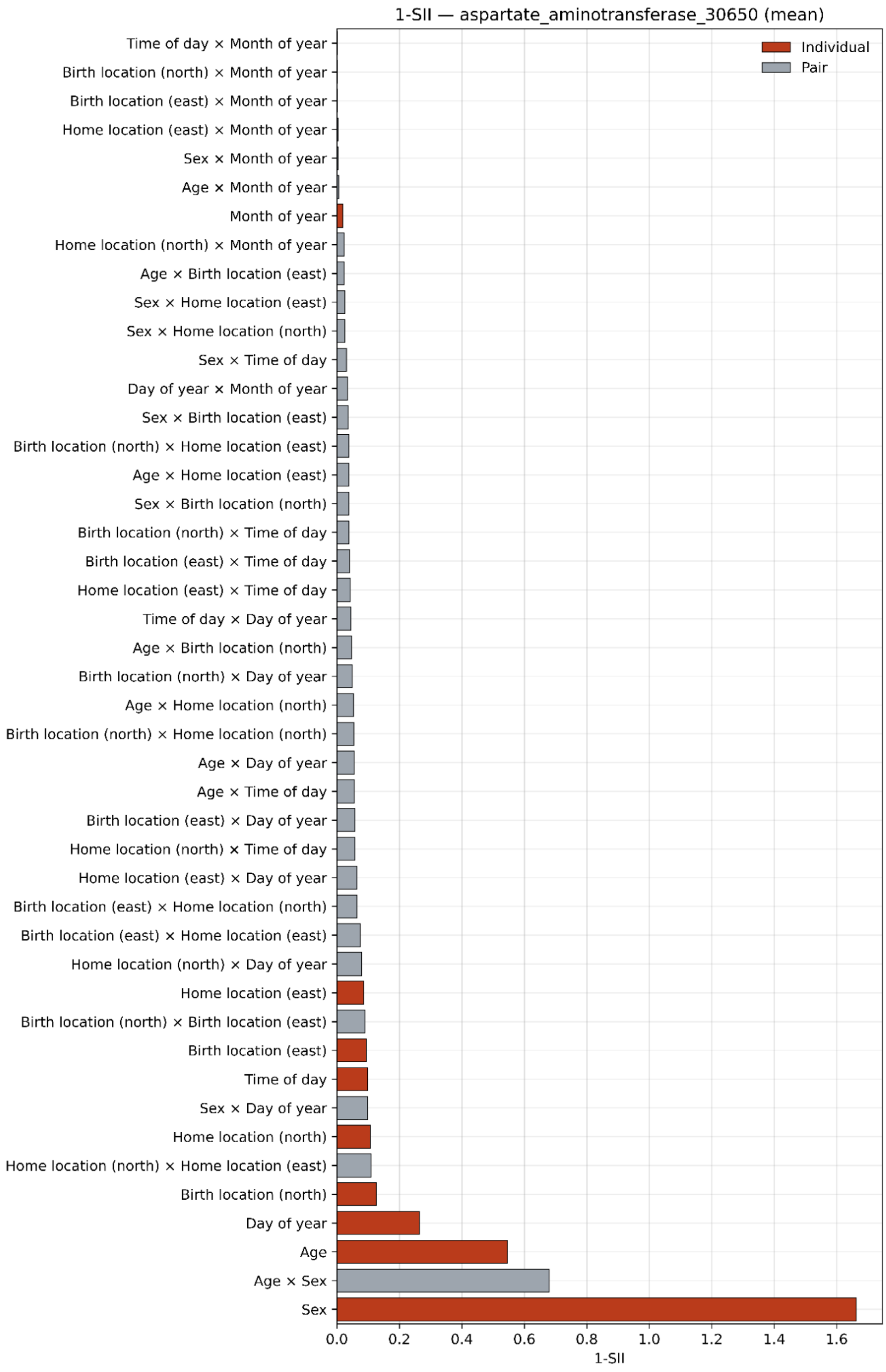

#### Supplementary Figure 4c

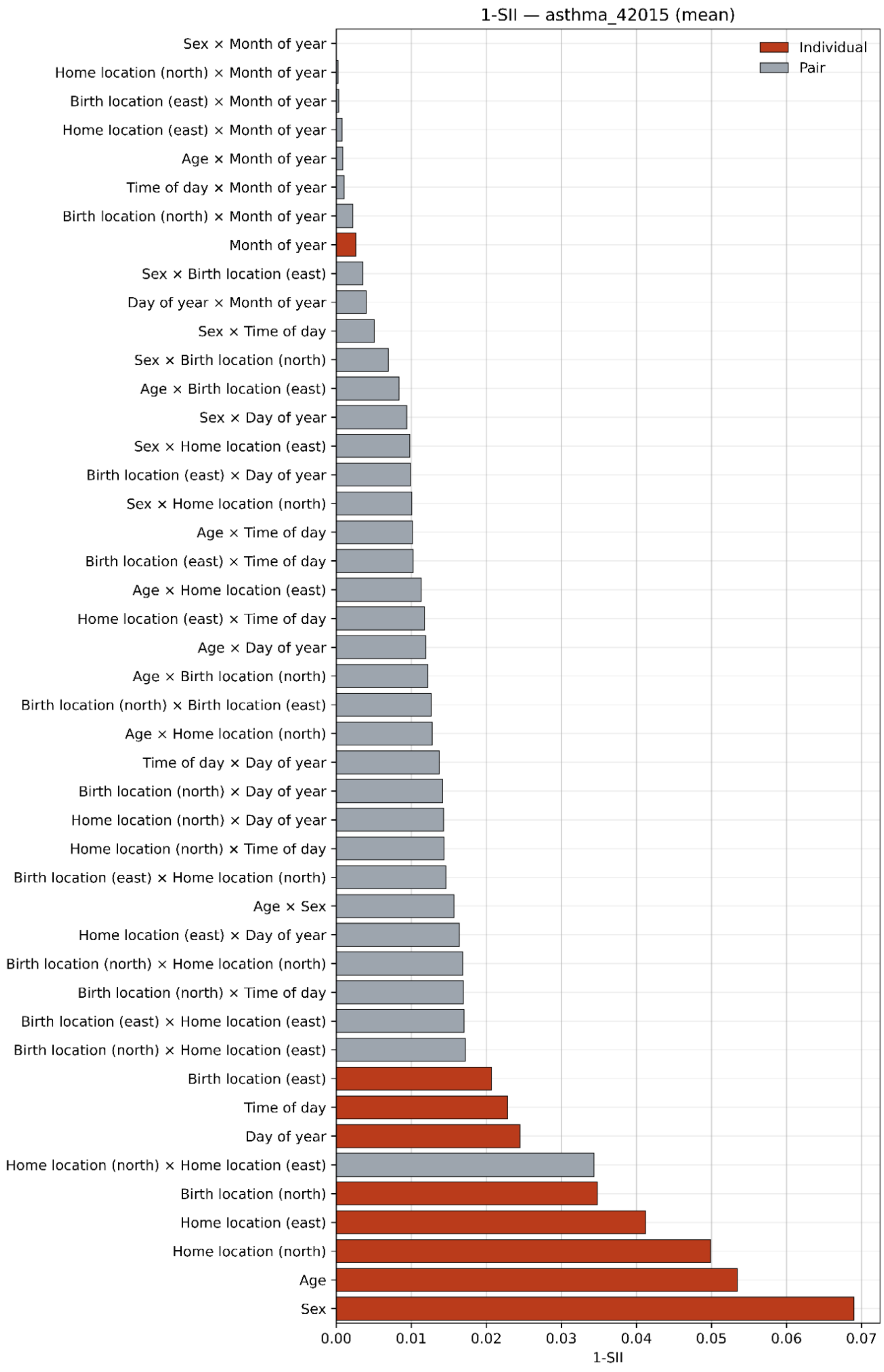

#### Supplementary Figure 4d

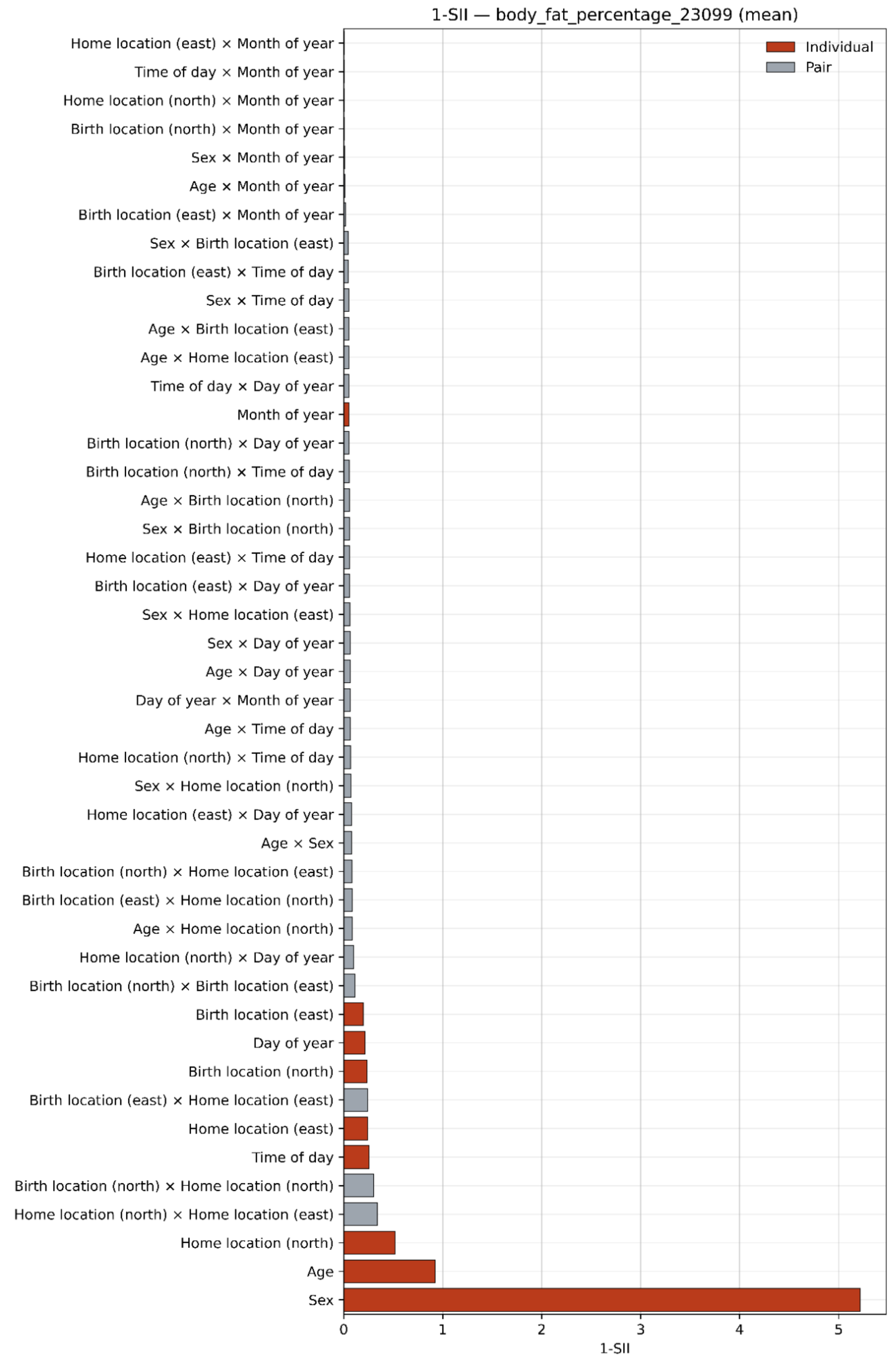

Supplementary Figure 4e

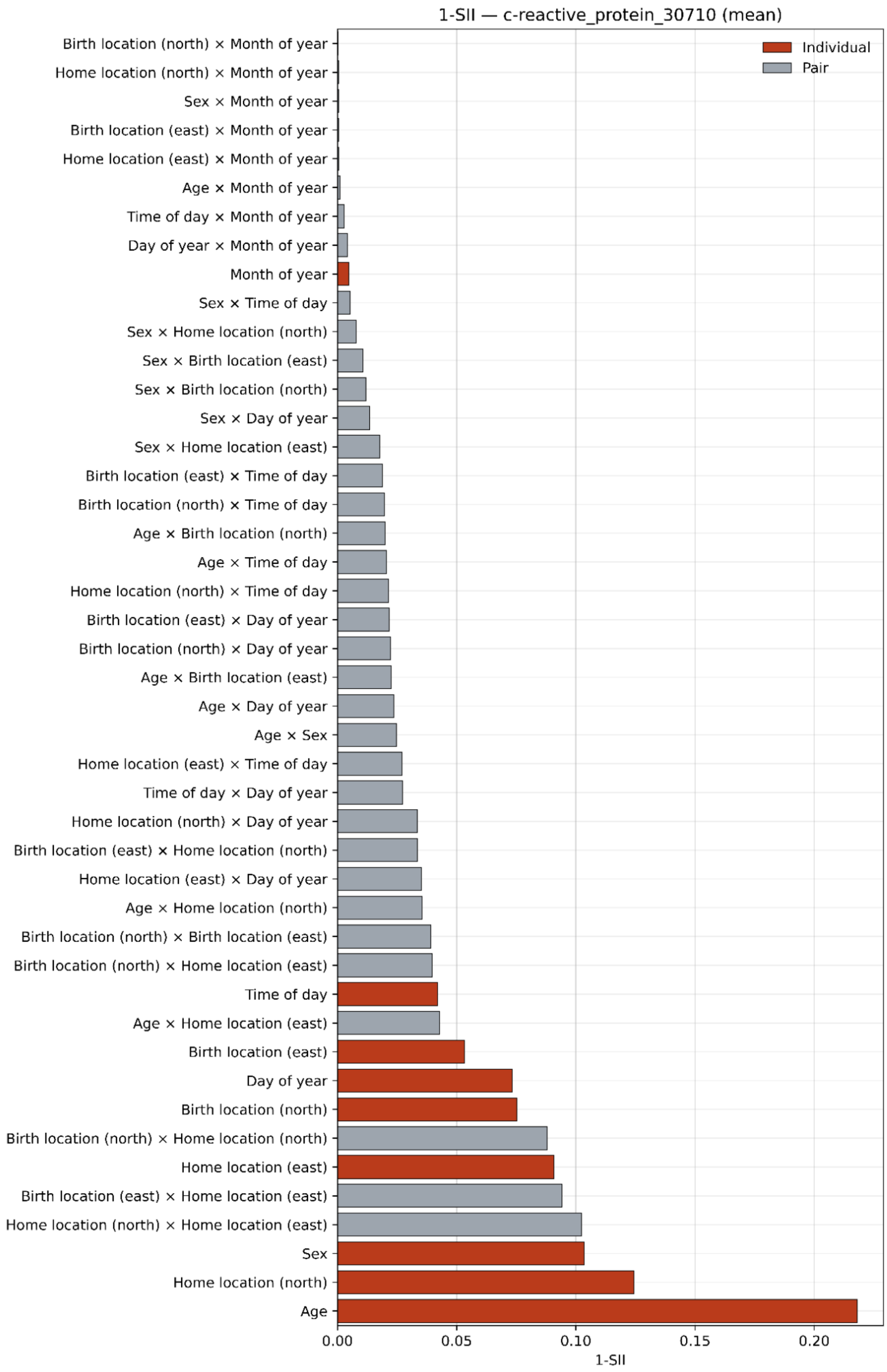

Supplementary Figure 4f

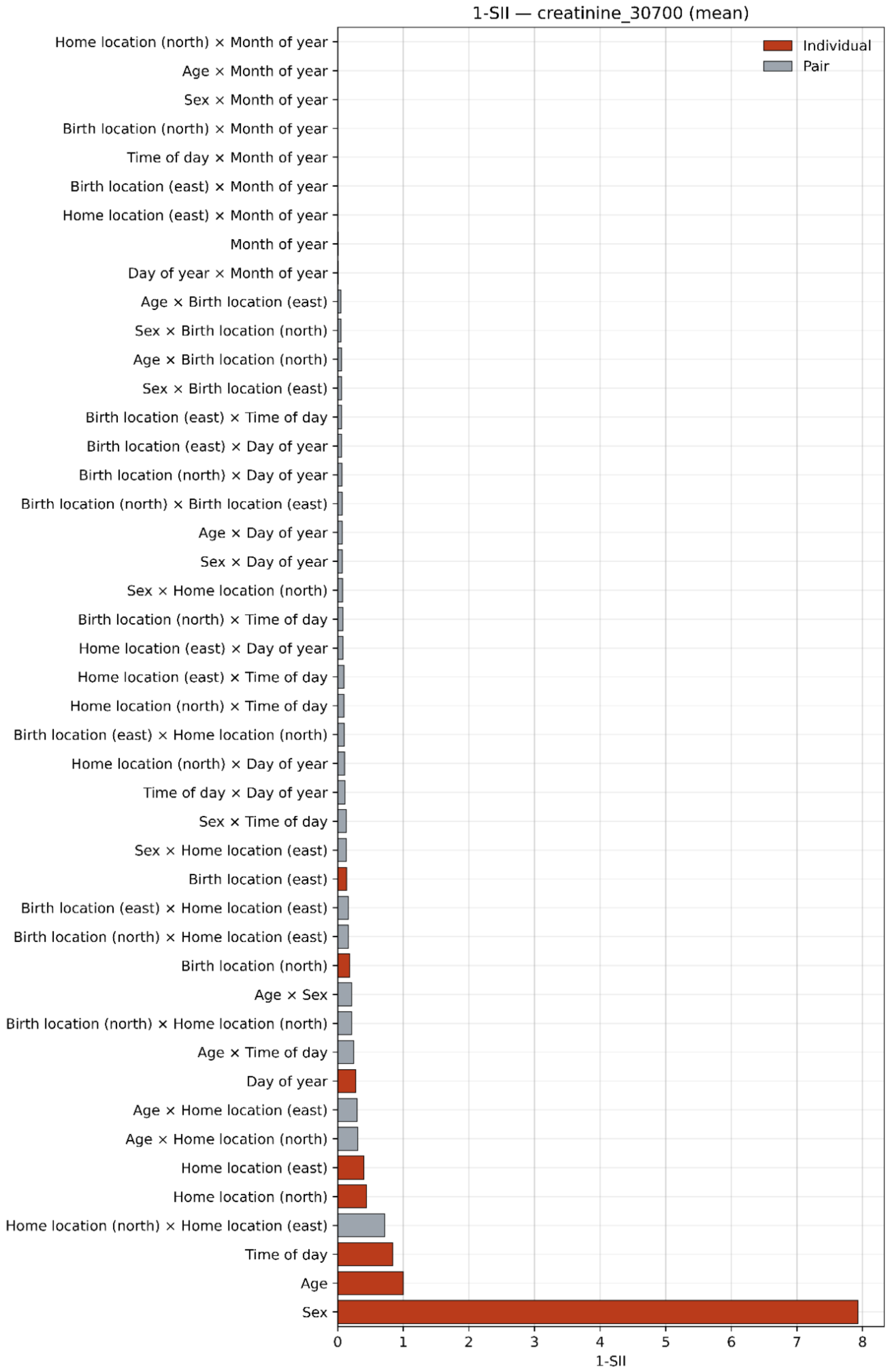

Supplementary Figure 4g

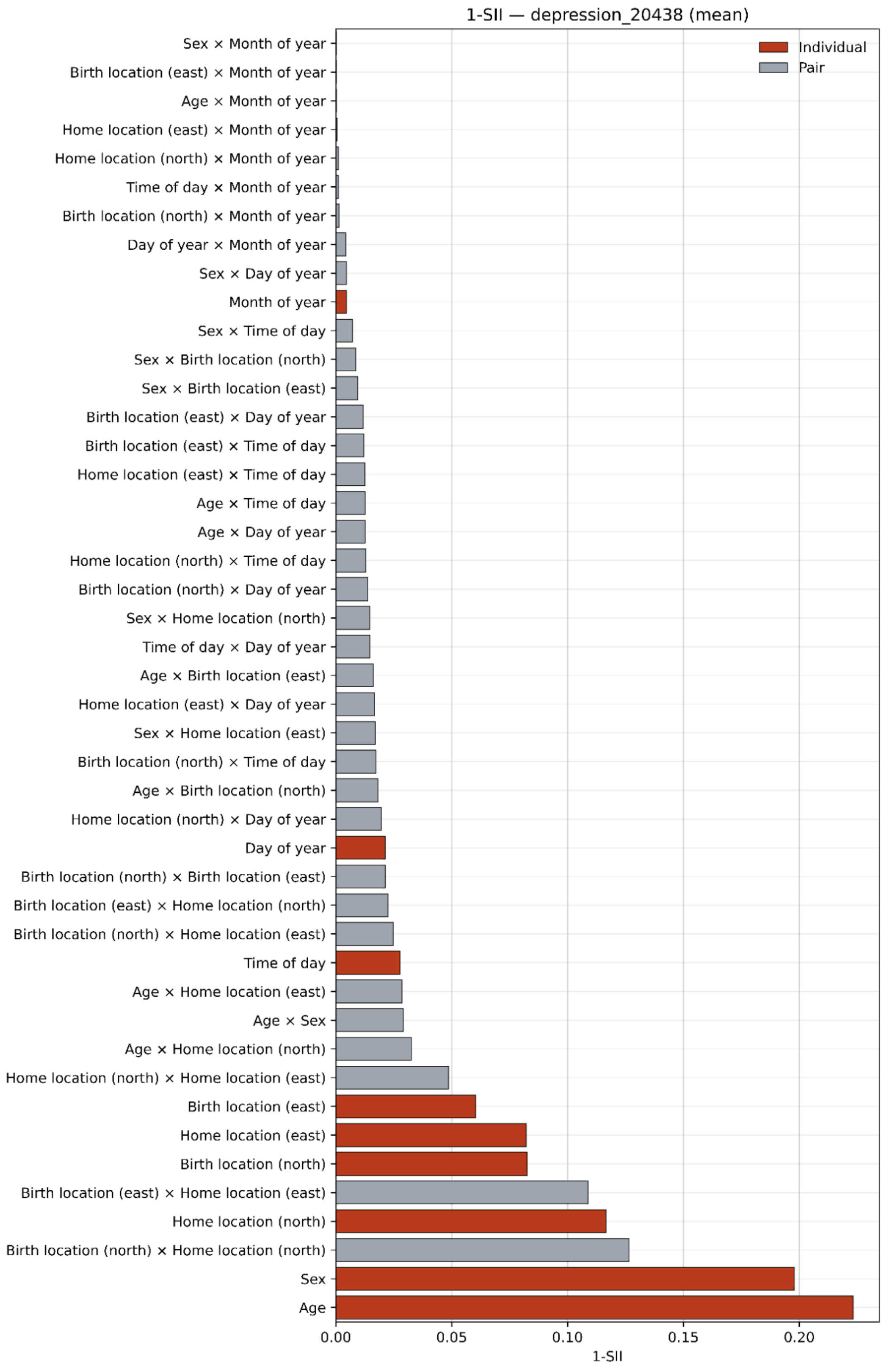

Supplementary Figure 4h

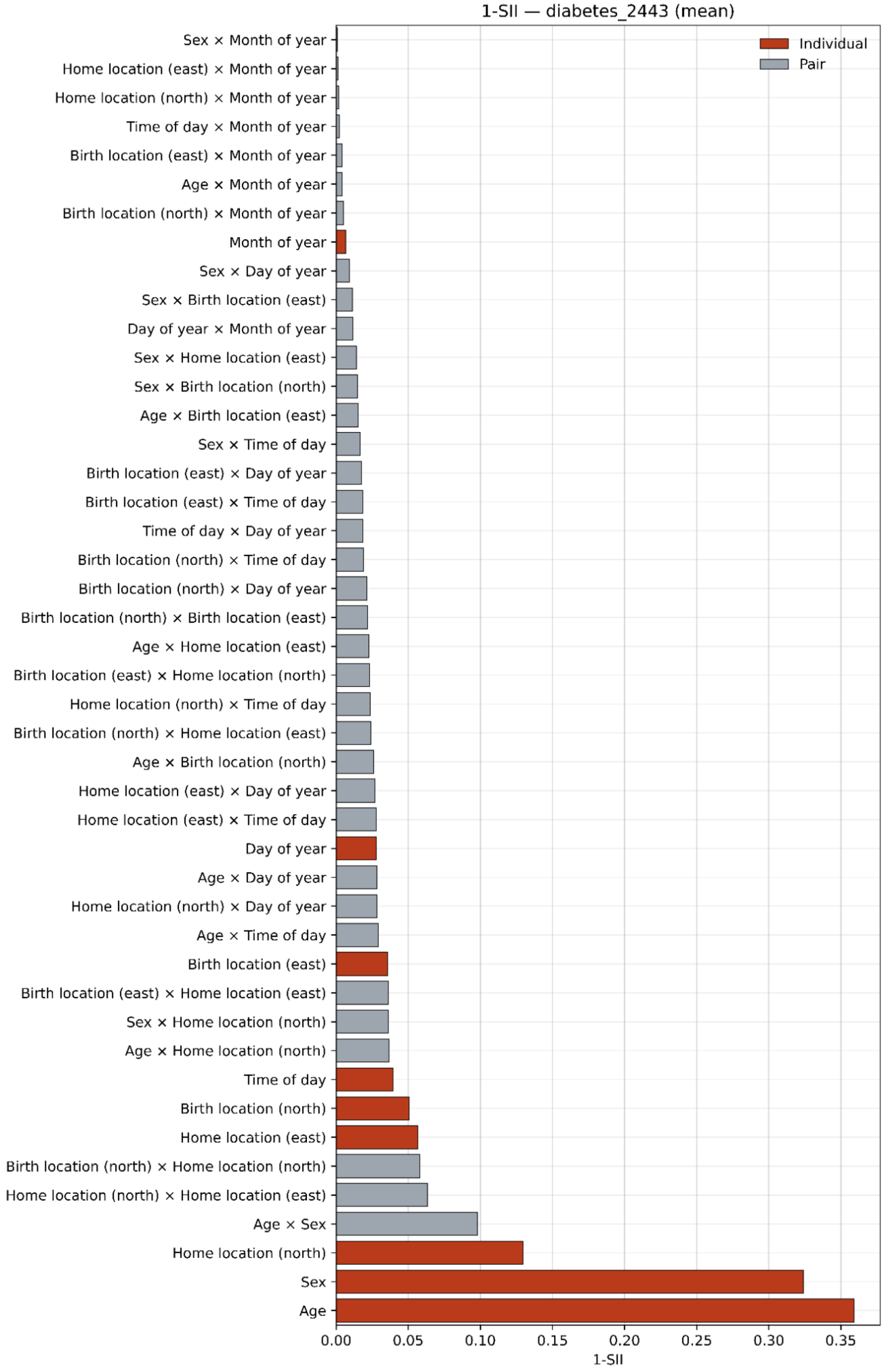

#### Supplementary Figure 4i

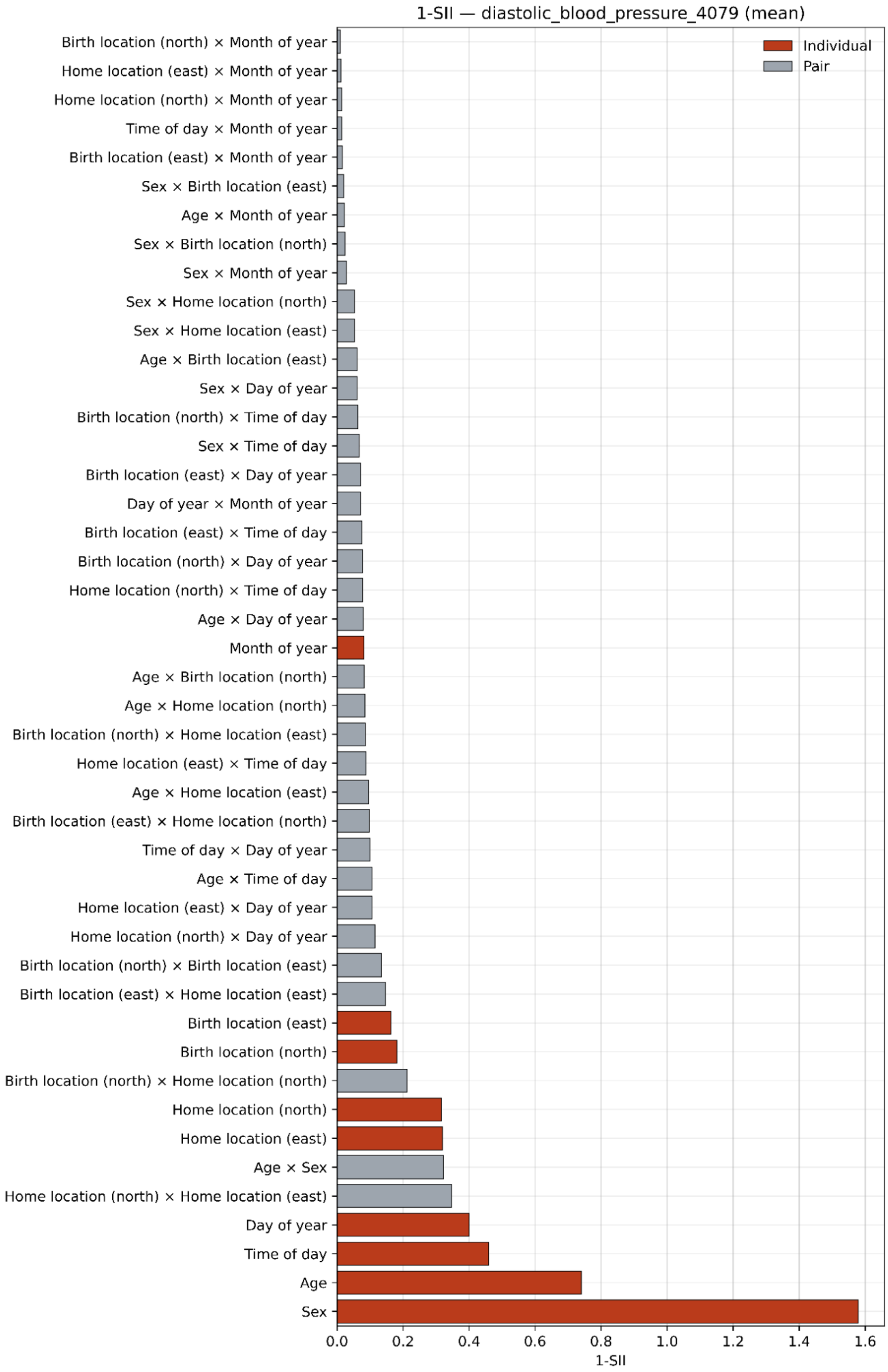

#### Supplementary Figure 4j

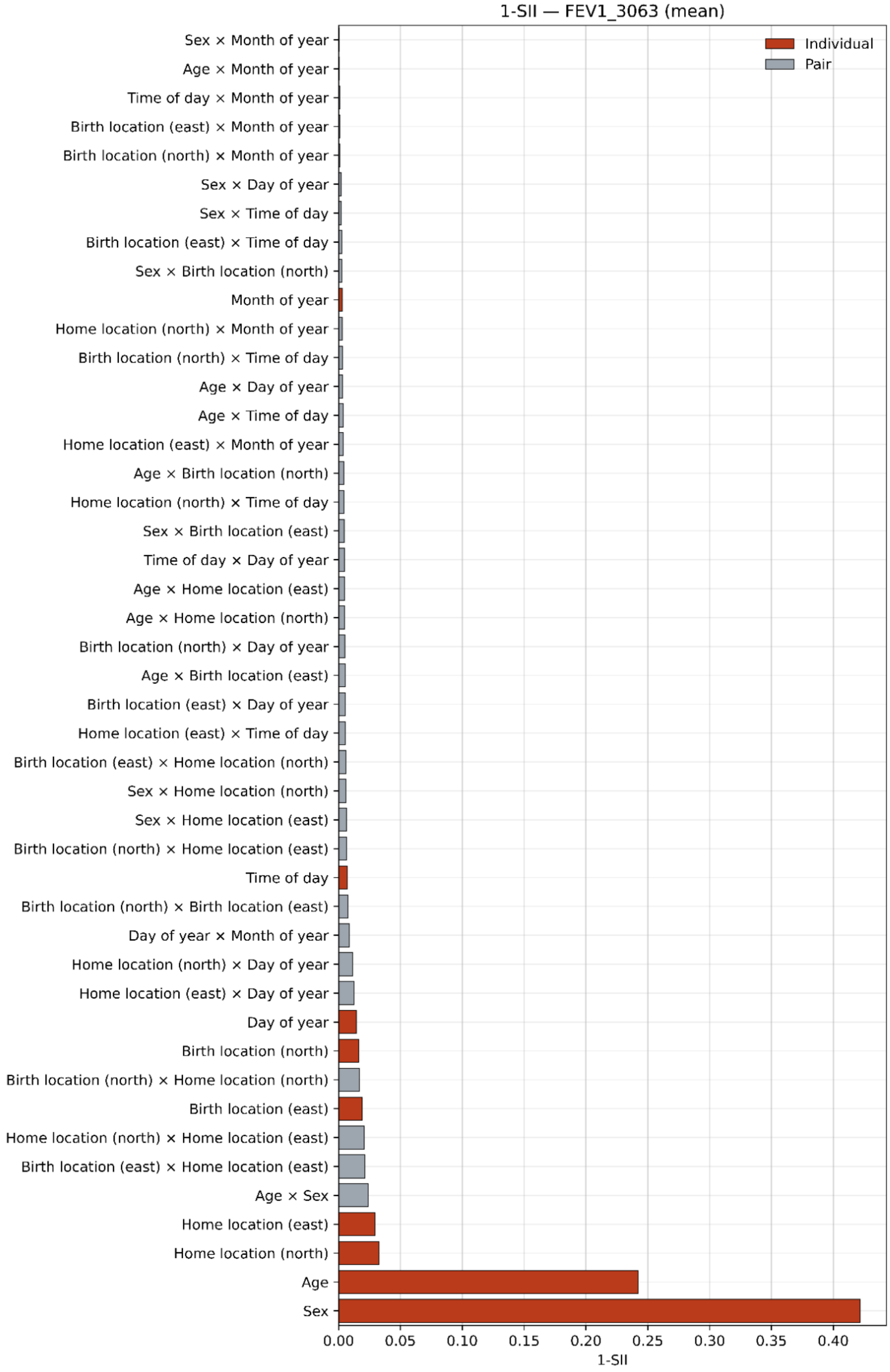

Supplementary Figure 4k

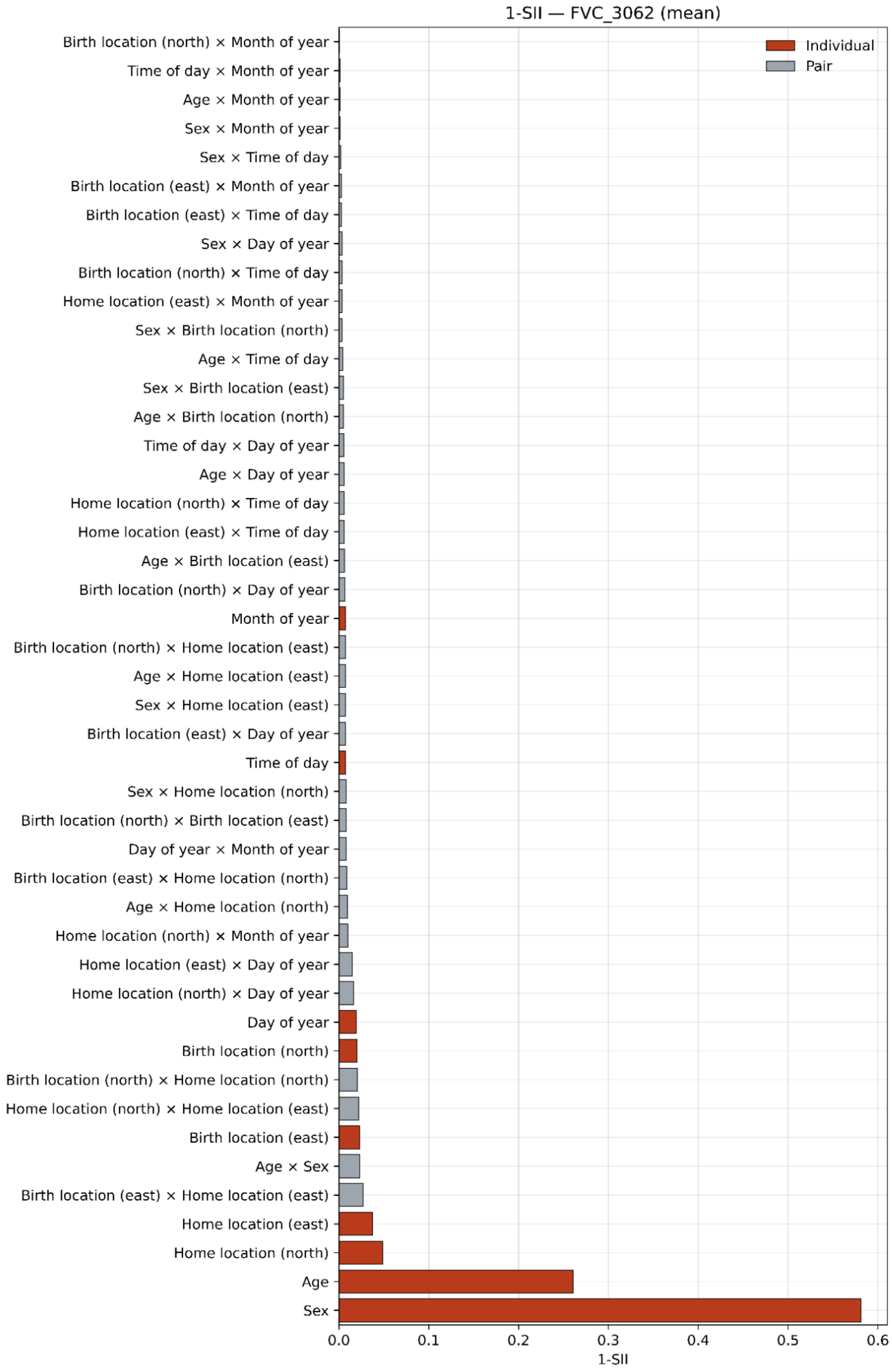

Supplementary Figure 4l

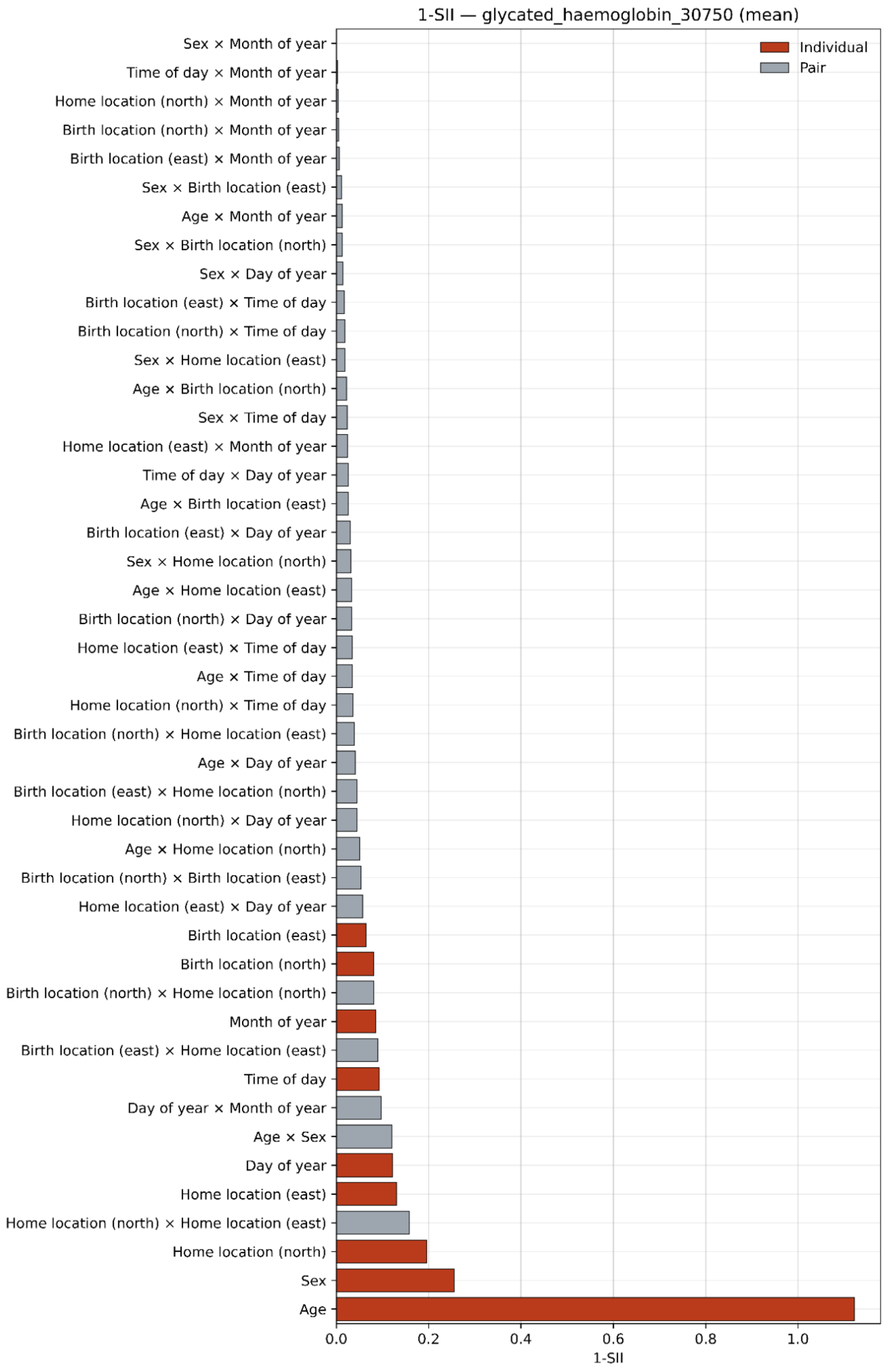

Supplementary Figure 4m

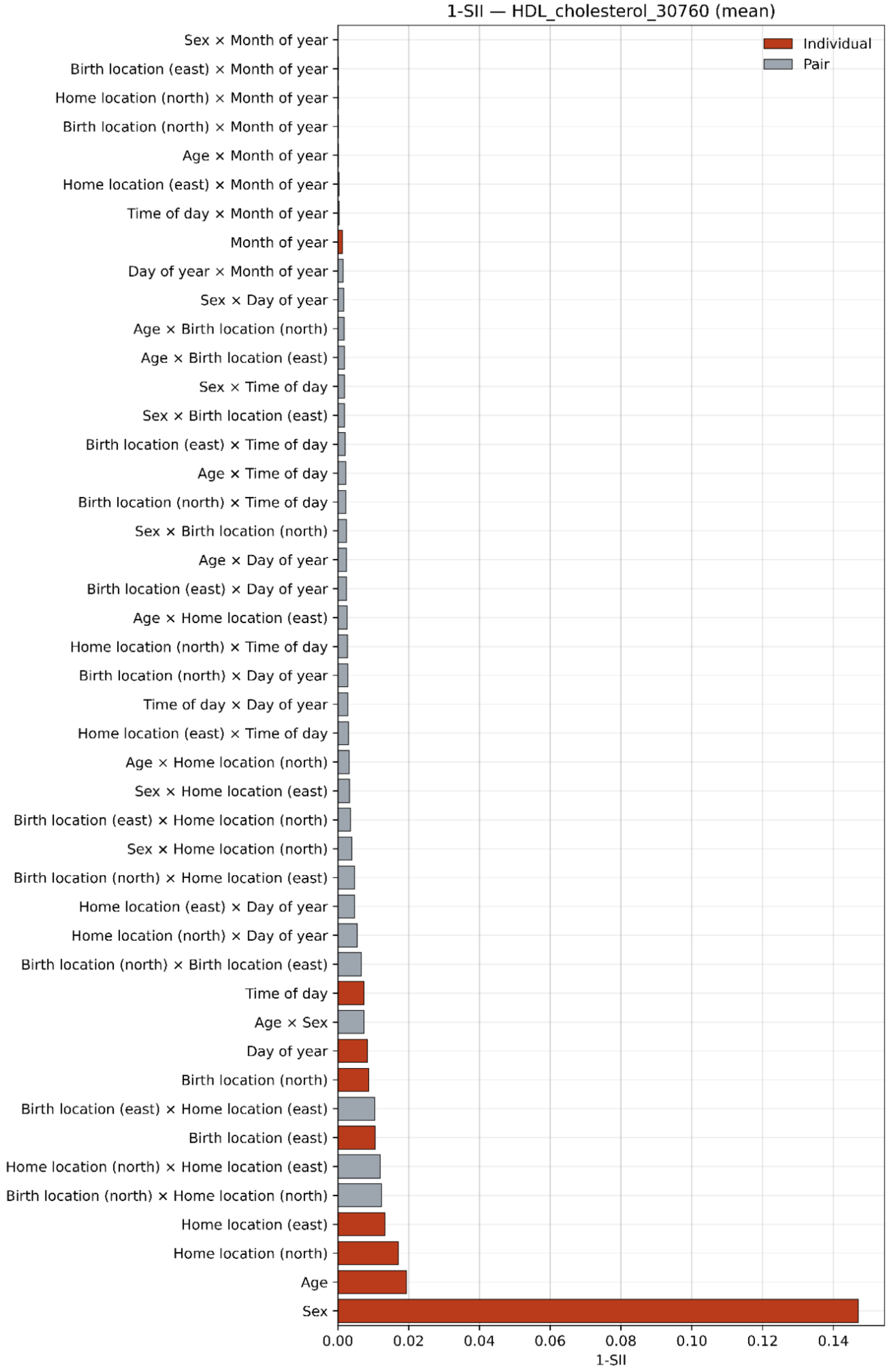

Supplementary Figure 4n

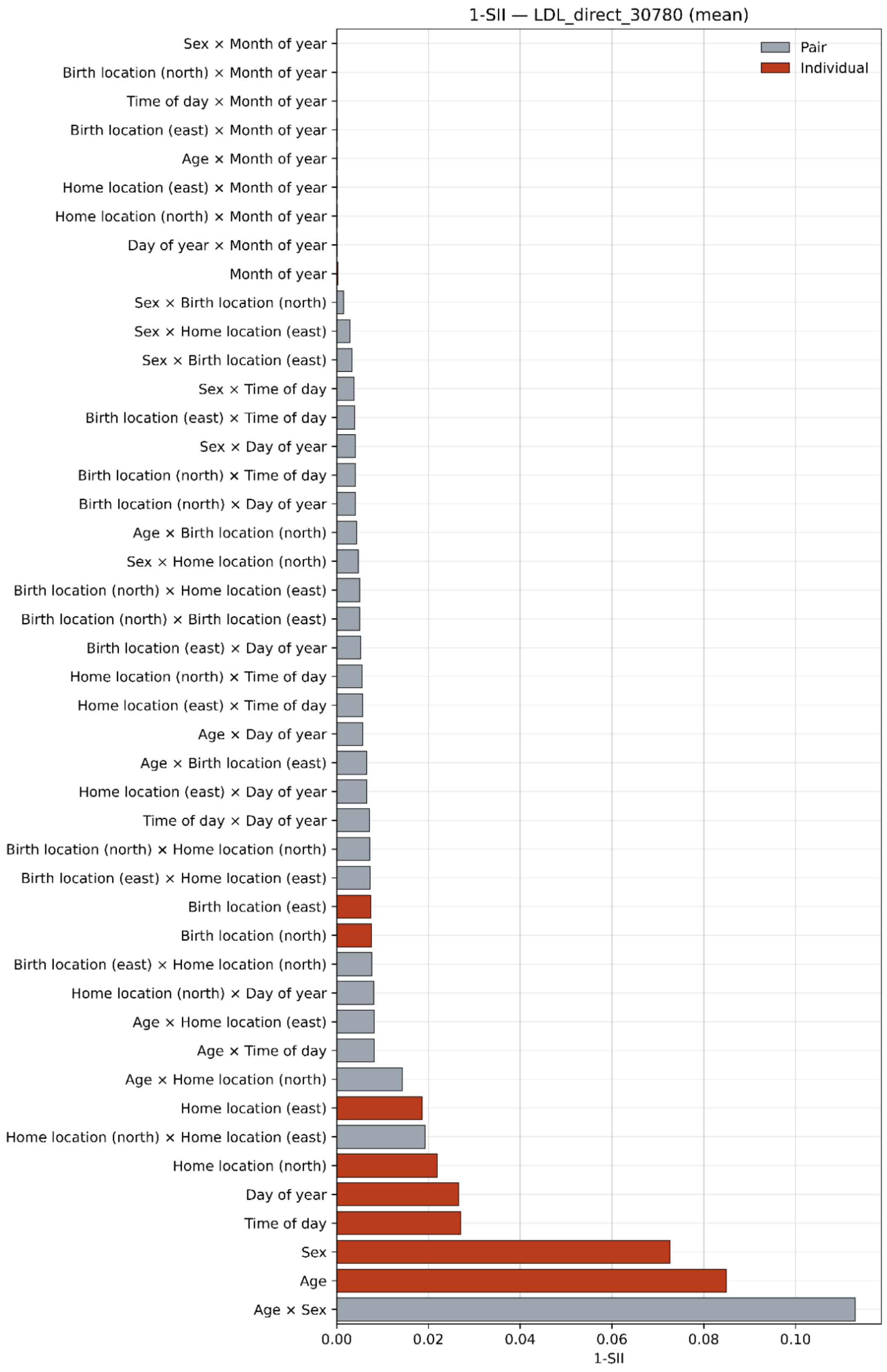

Supplementary Figure 4o

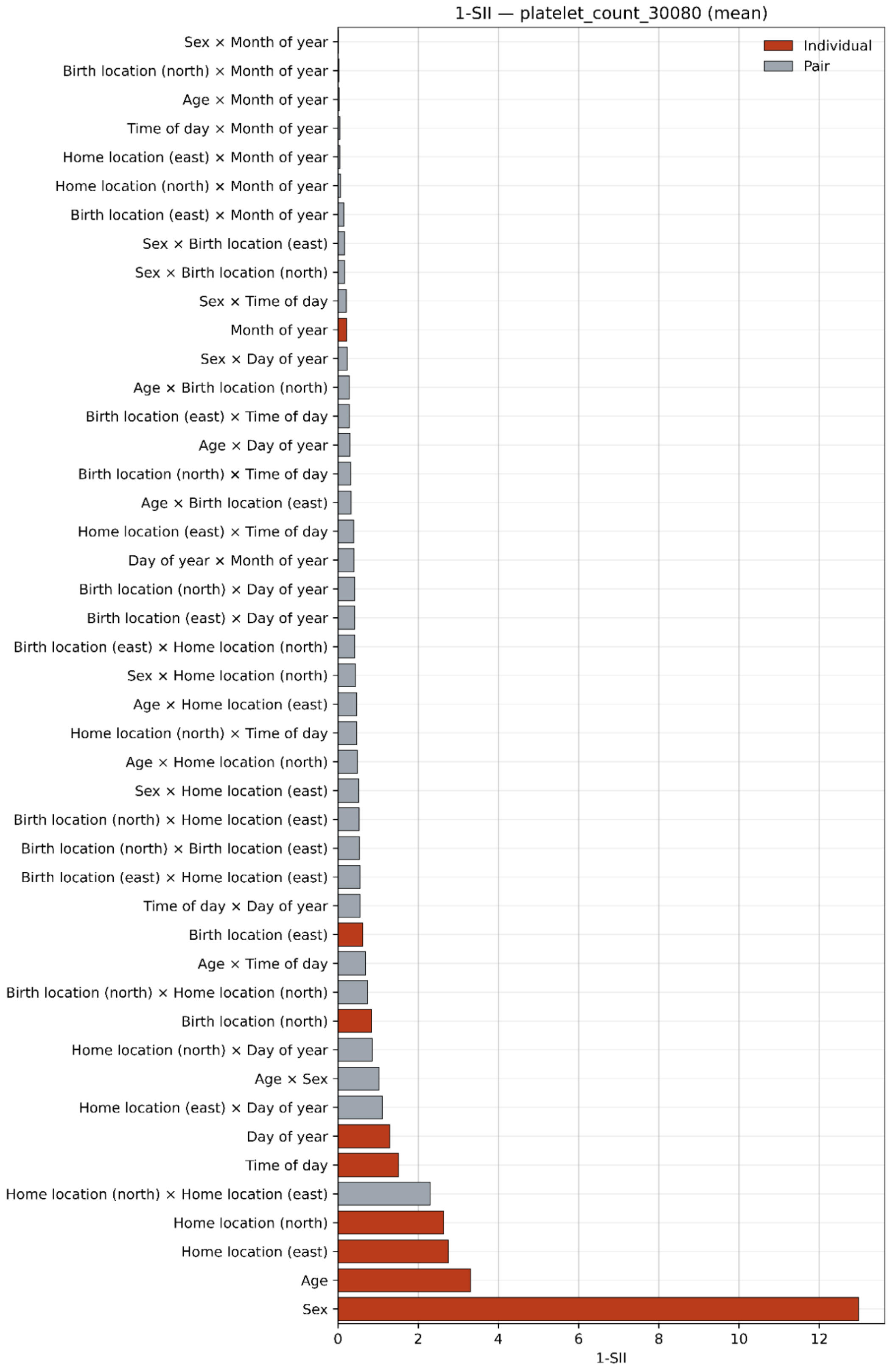

Supplementary Figure 4p

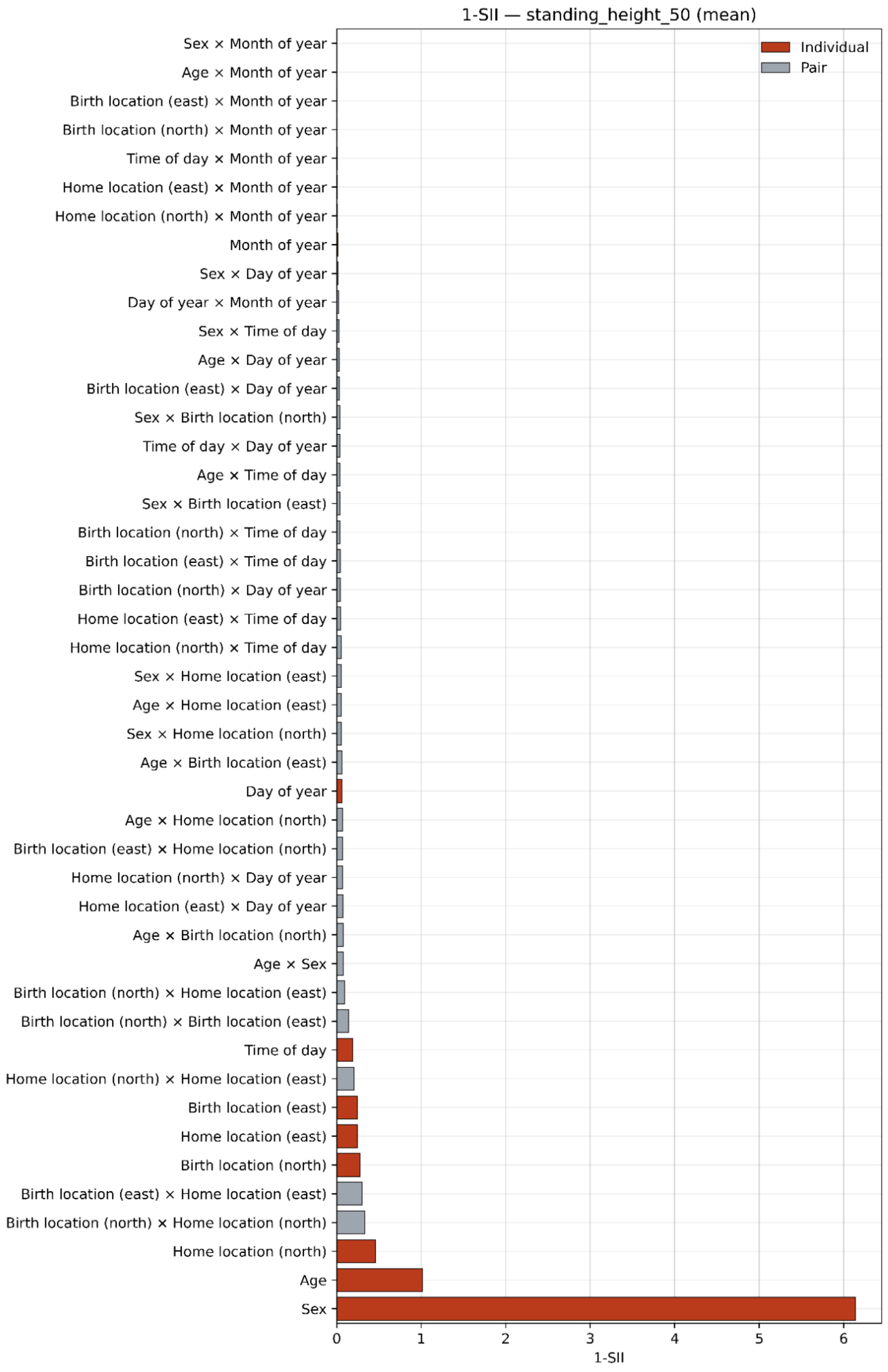

Supplementary Figure 4q

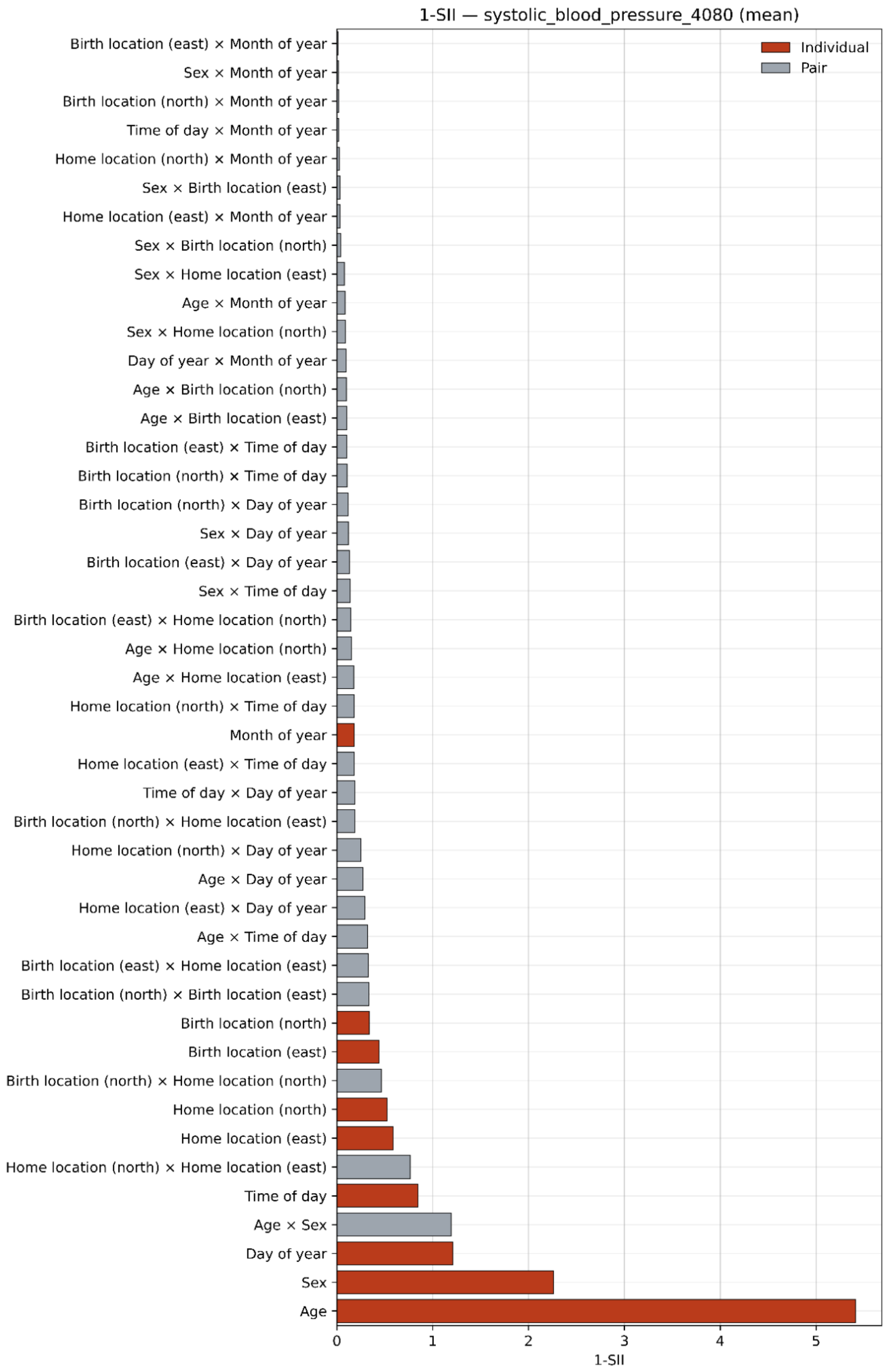

Supplementary Figure 4r

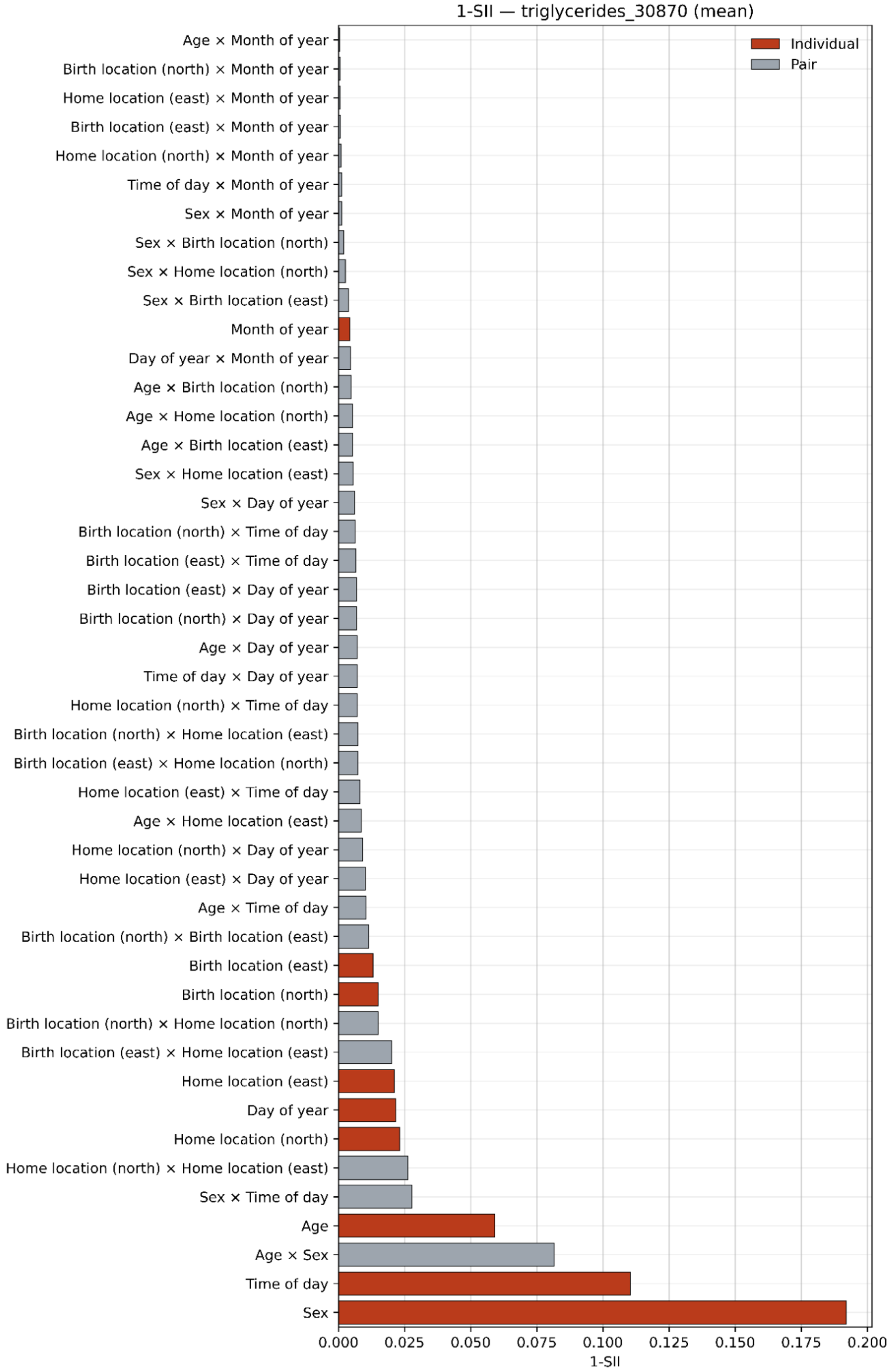

Supplementary Figure 4s

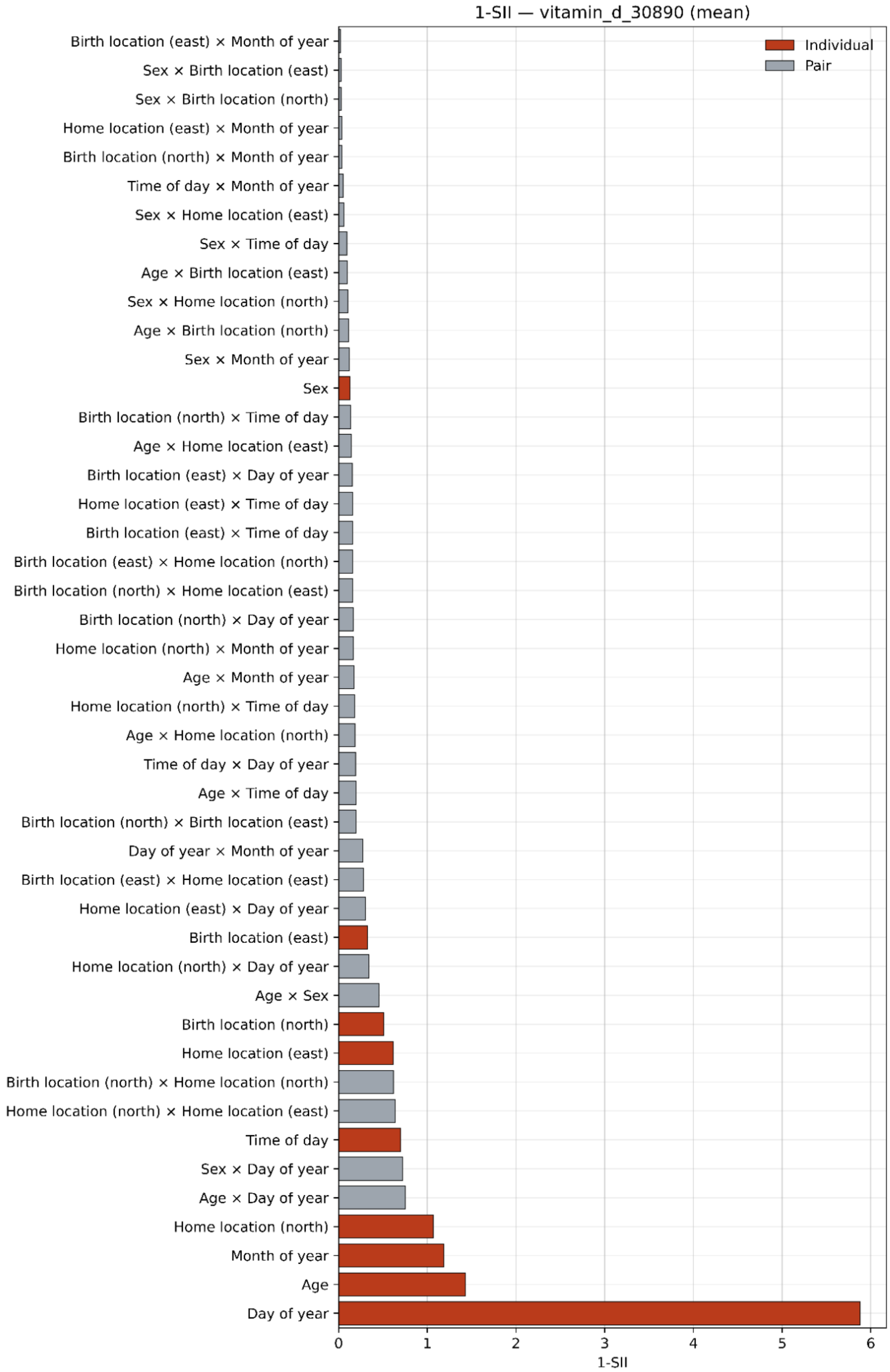

**Supplementary Figure 4: First-order Shapley Interaction Index (1-SII) values for all phenotypes.** Bar plots show the mean magnitude of the 1-SII values across the test set for all individual covariate features and feature pairs. Values for individual covariates are shown in red while pairs are in gray.

**(a) Alanine aminotransferase (ALT)**
**(b) Aspartate aminotransferase (AST)**
**(c) Asthma**
**(d) Body fat percentage**
**(e) C-reactive protein (CRP)**
**(f) Creatinine**
**(g) Depression**
**(h) Diabetes**
**(i) Diastolic blood pressure**
**(j) Forced expiratory volume in 1 second (FEV1)**
**(k) Forced vital capacity (FVC)**
**(l) Glycated haemoglobin**
**(m) HDL cholesterol**
**(n) LDL (cholesterol) direct**
**(o) Platelet count**
**(p) Standing height**
**(q) Systolic blood pressure**
**(r) Triglycerides**
**(s) Vitamin-D**
